# Pediatric and Adult Patients with ME/CFS following COVID-19: A Structured Approach to Diagnosis Using the Munich Berlin Symptom Questionnaire (MBSQ)

**DOI:** 10.1101/2023.08.23.23293081

**Authors:** Laura C. Peo, Katharina Wiehler, Johannes Paulick, Katrin Gerrer, Ariane Leone, Anja Viereck, Matthias Haegele, Silvia Stojanov, Cordula Warlitz, Silvia Augustin, Martin Alberer, Daniel B. R. Hattesohl, Laura Froehlich, Carmen Scheibenbogen, Lorenz Mihatsch, Rafael Pricoco, Uta Behrends

## Abstract

**Purpose:** A subset of patients with post-COVID-19 condition (PCC) fulfill the clinical criteria of myalgic encephalomyelitis / chronic fatigue syndrome (ME/CFS). To establish the diagnosis of ME/CFS for clinical and research purposes, comprehensive scores have to be evaluated.

**Methods:** We developed the Munich Berlin Symptom Questionnaires (MBSQs) and supplementary scoring sheets (SSSs) to allow for a rapid evaluation of common ME/CFS case definitions. The MBSQs were applied to young patients with chronic fatigue and post-exertional malaise (PEM) who presented to the MRI Chronic Fatigue Center for Young People (MCFC). Trials were retrospectively registered (NCT05778006, **NCT05638724).**

**Results:** Using the MBSQs and SSSs, we report on ten patients aged 11 to 25 years diagnosed with ME/CFS after asymptomatic SARS-CoV-2 infection or mild to moderate COVID-19. Results from their MBSQs and from well-established patient-reported outcome measures indicated severe impairments of daily activities and health-related quality of life.

**Conclusions:** ME/CFS can follow SARS-CoV-2 infection in patients younger than 18 years, rendering structured diagnostic approaches most relevant for pediatric PCC clinics. The MBSQs and SSSs represent novel diagnostic tools that can facilitate the diagnosis of ME/CFS in children, adolescents, and adults with PCC and other post-viral syndromes.

**What is known:** ME/CFS is a frequent debilitating illness. For diagnosis, an extensive differential diagnostic workup is required and the evaluation of clinical ME/CFS criteria. ME/CFS following COVID-19 has been reported in adults but not in pediatric patients younger than 19 years of age.

**What is new:** We present novel questionnairs (MBSQs), as tools to assess common ME/CFS case definitions in pediatric and adult patients with post-COVID-19 condition and beyond. We report on ten patients aged 11 to 25 years diagnosed with ME/CFS following asymptomatic SARS-CoV-2 infection or mild to moderate COVID-19.

## INTRODUCTION

The coronavirus disease 2019 (COVID-19) pandemic and its long-term-sequelae elicited an unprecedented healthcare crisis worldwide. Beyond acute morbidity and mortality due to infections with severe acute respiratory coronavirus type 2 (SARS-CoV-2) [1; 2], a plethora of post-acute sequelae of COVID-19 (PASC) (often referred to as Long COVID) with or without major organ damage due to SARS-Cov-2 infection are contributing to the post-pandemic burden and are increasingly challenging healthcare systems and societies [3-7].

Most individuals recover from PASC within a few months, but some develop a long-lasting disorder that can severely impair daily function, participation, and health-related quality of life (HRQoL) [4; 8-10]. A post-COVID-19 condition (PCC) (ICD-10 U09.9!) was defined by the World Health Organization (WHO) as continuing or new development of symptoms three months (children: within three months) after the initial SARS-CoV-2 infection, lasting for at least two months and not explained otherwise [11; 12].

PASC were estimated to affect at least 65 million individuals worldwide, with a prevalence of about 10% infected cases and a lower prevalence in children compared to adolescents and adults [4; 13]. Estimating pediatric PCC prevalence is still challenging [8; 14], with 0.8 to 13% reported in controlled cohorts [15; 16] and 2.0 to 3.5% calculated in a meta-analysis covering initially non-hospitalized children and adolescents [17].

COVID-19 sequelae may manifest with a wide variety of symptoms, including fatigue, shortness of breath, cognitive dysfunction, pain, sleep disorder, and/or mood symptoms. These symptoms can persist, fluctuate, or relapse and may have a significant impact on everyday functioning [11; 12; 18-20]. Some patients suffer from exertion intolerance with a worsening of symptoms after mild physical and/or mental activities, known as post-exertional malaise (PEM) [21; 22]. PEM can last for days or weeks and is recognized as a cardinal symptom of myalgic encephalomyelitis / chronic fatigue syndrome (ME/CFS) [23; 24]. ME/CFS following COVID-19 has been reported in adults [21; 25; 26] and in a 19-year-old male from the U.S. [27; 28] but, to our knowledge, not yet in younger patients. However, overlapping symptoms of PACS and ME/CFS have been described in pediatric patients [29].

ME/CFS is a complex, chronic neurological disorder (ICD-10 G93.3), triggered mostly by infections and rarely by non-infectious life events [30-32]. Core symptoms include reduced daily functioning with fatigue not alleviated by rest, PEM usually lasting more than a day, unrefreshing sleep, neurocognitive deficits (“brain fog”) and/or orthostatic intolerance (OI), with additional symptoms in most cases [33]. Hypothesized pathogenic mechanisms of PCC and ME/CFS overlap, including viral persistence, latent virus reactivation, inflammation, autoimmunity, endothelial dysfunction, and microbiome dysbiosis [23; 34]. Common risk factors of PCC and ME/CFS are female gender, late adolescence or early adulthood, as well as pre-existing chronic health issues [14; 21; 31; 35; 36].

Population-based, pre-pandemic estimates of ME/CFS prevalence ranged from 0.1% to 0.89% in adults [37-40] and from 0.75% to 0.98% in adolescents and children [41; 42], with a high number of undetected cases [42]. Current estimates predicted at least a doubling of ME/CFS cases due to severe PCC [21; 27; 34; 43; 44]. A total of 350.000 and 400.000 ME/CFS cases were documented in 2018 and 2019, and almost 500.000 cases in 2021 by the German National Association of Statutory Health Insurance Physicians (KBV) [45], with no data for children and adolescents available yet.

Since up to now, no reliable diagnostic biomarker for ME/CFS was established, complex disorders with compromising chronic fatigue require a thorough differential diagnostic workup and an evaluation of clinical ME/CFS criteria. Most commonly used are the Canadian consensus criteria (CCC) [46] and the broader criteria established by the former Institute of Medicine (IOM) to define “systemic exertion intolerance disease (SEID)” [47]. For children and adolescents, the CCC were adapted by a “pediatric case definition” of L.A. Jason and colleagues (abbreviated here as PCD-J) [48] and in a less restrictive way by the “clinical diagnostic worksheet” designed by P.C. Rowe and colleagues (abbreviated here as CDW-R) [49]. A symptom duration of at least six months is usually required for adult patients [31]. It was recently suggested to be reduced cross-age to facilitate early treatment by the National Institute of Health and Care Excellence (NICE) [50]. For children and adolescents, the required disease duration is three months in the CCC as well as the IOM and the PCD-J [46-48].

ME/CFS care requires a holistic, longitudinal approach, including extensive patient education, the palliation of symptoms, and adequate psychosocial support. Patients must be carefully guided in “pacing” strategies to avoid PEM (“crashes”) [50]. Since graded-exercise strategies (GET) can be harmful to patients with PEM [51] they should not be recommended for patients with ME/CFS [50], although they may be potentially helpful in other forms of PCC [52],

Early identification of patients with ME/CFS is crucial in order to avoid mismanagement and secondary damage, including suicidality. With adequate care, ME/CFS can improve in a substantial number of patients, with recovery documented for the majority of affected children and adolescents within ten years [32]. However, recovery does not imply absence of functional impairment [53]. The lack of ME/CFS-specific knowledge among healthcare professionals [54-56], together with an increasing ME/CFS prevalence, renders more patients at risk of insufficient care and secondary disease.

A challenge in clinical care and research for ME/CFS is the use of various diagnostic criteria and the lack of specific symptoms. To increase diagnostic sensitivity, the frequency and severity of symptoms should be assessed [57; 58].

With the Munich Berlin Symptom Questionnaires (MBSQs) we aimed to facilitate the diagnostic approach to adults, adolescents, and children with chronic fatigue following COVID-19 and beyond. The MBSQs represent novel tools for an age-adapted, standardized evaluation of the most common sets of clinical ME/CFS criteria in clinical and research settings.

Here, we present bi-lingual versions of the MBSQs and report on the first ten patients diagnosed with PCC and ME/CFS using the MBSQ in structured medical interviews at our MRI Chronic Fatigue Center For Young People (MCFC). Our Post-COVID clinic was implemented within the MCFC as part of the “Post-COVID Kids Bavaria” project to provide pediatric care and research in the context of severe COVID-19 sequelae [59].

## PATIENTS AND METHODS

### Inclusion Criteria and Clinical Assessment

Ten patients were diagnosed at the MCFC with PCC and ME/CFS using the German versions of the novel MBSQs and the supplementary scoring sheets (SSSs) (can be requested from authors, for English translation see **Supplementary Materia**) (see full description of the MBSQs below). The collection and publication of medical data from these patients was approved by the TUM Ethics Committee (116/21, 511/21). Written informed consent was obtained from all participants (or parents) prior to inclusion. All patients sought care at the MCFC with a history of confirmed (positive reverse transcription polymerase chain reaction (RT-PCR)) or probable (anti-SARS-CoV-2 IgG without prior COVID-19 vaccination and typical COVID-19 symptoms) SARS-CoV-2 infection and with post-viral symptoms lasting for more than three months, in accordance with WHO definitions of the PCC at any age.

Before visiting the MCFC, the patients had been asked to complete various questionnaires in a stepped routine process, including well-established patient-reported outcome measures (PROMs) to assess fatigue (Fatigue Severity Scale (FSS) [60] or Chalder Fatigue Scale (CFQ) [61]), PEM (DePaul Symptom Questionnaire - Post-Exertional Malaise (DSQ-PEM)) [24], limitations in daily functioning (Bell Score) [62], and HRQoL during the last four weeks (Short Form-36 Health Survey (SF-36)) [63]. The MBSQ was developed in 2020 and was provided together with other questionnaires prior to the personal visit to the MCFC.

The Bell Score measures daily functioning on a scale from 0% to 100%, with 100% representing normal daily functioning [62]. The DSQ-PEM provides a Likert scale for the frequency (0-4) and severity (0-4) of five different PEM-related symptoms and evaluates the duration of PEM [24]. The SF-36 consists of eight dimensions, including i) physical functioning, ii) social functioning, iii) vitality, iv) general health, v) mental health, vi) role physical, vii) role emotional, and viii) bodily pain. The score of each dimension is scaled to 0 – 100, with 0 representing the worst and 100 the best health status. The MCFC is focussing on patients with significant fatigue, indicated by a mean score of ≥ 5 (maximum: 7) in the FSS [64] or of ≥ 4 (maximum: 11) in the CFQ bimodal score [65] together with long-lasting PEM (≥14 hours) and reduced daily functioning.

Laboratory and technical investigations were performed at the MCFC and/or prior to admission to exclude other disorders that might explain these symptoms. The panel of analyses was selected depending on the individual symptoms, with a core set of routine analyses in line with prior suggestions [49]. Routine blood analyses included a differential cell count as well as C-reactive protein (CRP), liver, kidney, and thyroid function parameters, HbA1c, total serum immunoglobulins, antinuclear antibodies (ANA), antibodies against thyroid peroxidase (TPO), morning cortisol, antibodies against SARS-CoV-2 and Epstein-Barr virus (EBV), and EBV DNA load in blood and throat washes (PCR), supplemented by analyses of urine and stool (calprotectin, blood). Routine technical investigations included pulmonary function testing (PFT), electrocardiography (ECG), and ultrasound cardiography (UCG). If indicated, electroencephalography (EEG), cardiac or brain magnetic resonance tomography (MRT), ophthalmological and/or rheumatological assessments were added.

At the MCFC, patients were seen simultaneously by a pediatrician and psychologist or child and adolescent psychiatrist, each trained in ME/CFS, in order to provide a thorough diagnostic assessment. The pediatrician performed a comprehensive interview and physical examination, the psychologist or psychiatrist carried out the psychological evaluation. To assess any OI, including a possible postural orthostatic tachycardia syndrome (PoTS) or orthostatic hypotension (OH), all patients underwent a 10-minute NASA lean test [66]: heart rate (HR) and blood pressure were measured every minute by an electronic device (Carescape V100 Vital Signs Monitor). Patients were asked to hold a supine position for five minutes, then to stand upright and motionless (leaning against the wall with the shoulders and placing the feet two to six inches away from the wall) for ten minutes, and finally to lie down again for five minutes. They were instructed to report any novel symptoms during the whole procedure [49]. The average HR while supine was defined as baseline, and PoTS was defined by a sustained HR ≥ 120 beats per minute (bpm) and/or an increase by HR ≥ 40 bpm for individuals ≤ 19 years and ≥ 30 bpm for individuals > 19 years, together with a history of orthostatic symptoms for at least three months [67; 68]. A psychological evaluation was either provided externally or by psychologists trained in ME/CFS at the MCFC.

To establish the ME/CFS diagnosis, clinical criteria were evaluated in semi-structured interviews. The MCFC physician went through the pre-filled MBSQ together with the patient (and his/her parents) to avoid any misunderstanding regarding the presence, frequency, and severity of symptoms as well as the duration of PEM. All answers addressing PEM were carefully re-evaluated, and patients were asked to provide examples of typical PEM triggers and “crashes”. The pre-filling procedure at home allowed for taking enough time to answer the questionnaire and enabled the physician to focus on aspects to be clarified during the clinical visit. The MBSQ provides additional space for physician’s notes next to each question in the MBSQ. After the visit, the physician filled in the SSS for the respective age group in line with answers consented to in the interview and assessed the various case definitions. A final ME/CFS diagnosis was established if at least one case definition was matched and no other explanation of symptoms arose from adequate diagnostic workup. Each case was discussed in an interdisciplinary ME/CFS board, involving several physicians with ME/CFS expertise.

### Development of the Munich Berlin Symptom Questionnaire

The MBSQs and SSSs were designed as a joint collaboration of ME/CFS experts from the MCFC in Munich, the Charité Fatigue Center (CFC) in Berlin, the German Association for ME/CFS in Hamburg, and the Research Center CATALPA in Hagen, Germany. They were developed for clinical and research purposes to facilitate the evaluation of diagnostic ME/CFS criteria in a comprehensive, semi-structured personal or telephone interview by physicians trained in ME/CFS.

We chose the IOM criteria and CCC, which require a disease duration of at least six months for adults (≥18 years) as recommended by the Centers for Disease Control and Prevention (CDC) [69] and the European Network for ME/CFS (EUROMENE) [31], respectively. To keep the MBSQs and SSSs for adults as short as possible, we designed a separate version for children and adolescents (< 18 years) that contained not only questions relevant to the CCC and IOM criteria but also additional questions to assess the PCD-J and CDW-R criteria. For practical reasons, the pediatric version of the MBSQs require three months of disease duration for any case definition, although the CDW-R originally suggested to provide only a preliminary diagnosis after three and a confirmed diagnosis after six months [49] (**Table 1**).

**Table 1.** MBSQ versions for different age groups.

| *Table 1. MBSQ versions for different age groups* |  |  |  |  |  |  |
| --- | --- | --- | --- | --- | --- | --- |
| **Version** | **Adressed period of symptoms** | **Age group** | **CCC** | **IOM** | **PCD-J** | **CDW-R** |
| **Children and Adolescents** | Past 3 months | 0-17 years | + | + | + | + |
| **Adults** | Past 6 months | ≥ 18 years | + | + | - | - |
| MBSQ, Munich Berlin Symptom Questionnaire;<br>CCC, Canadian Consensus Criteria, 2003 [46]; IOM, Criteria of the former Institute of Medicine, 2015 [47];<br>PCD-J, Pediatric Case Definition by Jason et al., 2006 [48] ; CDW-R, Clinical Diagnostic Worksheet by Rowe et al., 2017 [49]; |  |  |  |  |  |  |

We first developed the German versions of the MBSQs and SSSs. All English terms used to describe the symptoms in the original publications [46-49] were attributed to the most adequate of the eight symptom categories of the CCC (fatigue, PEM, sleep disorder, pain, neurocognitive, autonomic, neuroendocrine, and immunologic manifestations). Overlaps and differences were identified, and umbrella terms were introduced if necessary to provide a concise questionnaire. We aimed at the best match of all terms with terms in the original publications and adapted the wording, if necessary, during several rounds of clinical testing and discussion to optimize the understanding by patients and/or parents. To keep the MBSQ as concise as possible, the wording was not adapted for a better understanding for children. The MBSQ was not designed and not evaluated as a patient-reported outcome measure (PROM) and therefore is not recommended for use as a PROM. The final German versions were translated back to English, aiming at optimal consistency. The English versions was provided to only few English speaking patients at our centers none of which was included in this report.

As suggested in the DSQs by Jason and colleagues [57; 58], we used a 5-point Likert scale for quantifying the frequency and severity of symptoms. In line with the DSQs, the MBSQs require an at least moderate frequency and severity (≥2) to support the ME/CFS diagnosis. However, instead of two separate columns for “0” answers regarding severity and frequency in the DSQs, the MBSQs provide a single column to indicate that symptoms were not present. In the MBSQs, four additional questions were provided in a dichotomous format to evaluate the presence or absence of distinct features of fatigue or neurocognitive manifestations. Three further questions provided space for three open answers each to gain information on the prominent triggers of PEM, the main symptoms of PEM, and the most bothering symptoms of ME/CFS. In contrast to the very comprehensive DSQ-2 [58], the MBSQ focuses on ME/CFS symptoms only, omitting any further evaluation of medical history.

## RESULTS

We developed the MBSQs and SSSs in German (request from authors) and English (**Supplementary Material**) as novel tools for the clinical assessment of ME/CFS in the context of PCC and beyond. They address the most commonly recommended ME/CFS case definitions (CCC, IOM) and, in the versions for children and adolescents, two additional pediatric case definitions (CDW-R, PCD-J) (**Table 1**) to facilitate semi-structured, age-adapted approaches to diagnosis. Here, we apply the MBSQs and SSSs to patients with PCC and report on the first ten patients diagnosed with ME/CFS according to MBSQ results after a thorough diagnostic workup at our MCFC (**Tables 1 and 2**, **Figures 1 and 2**).

**Figure 1.**
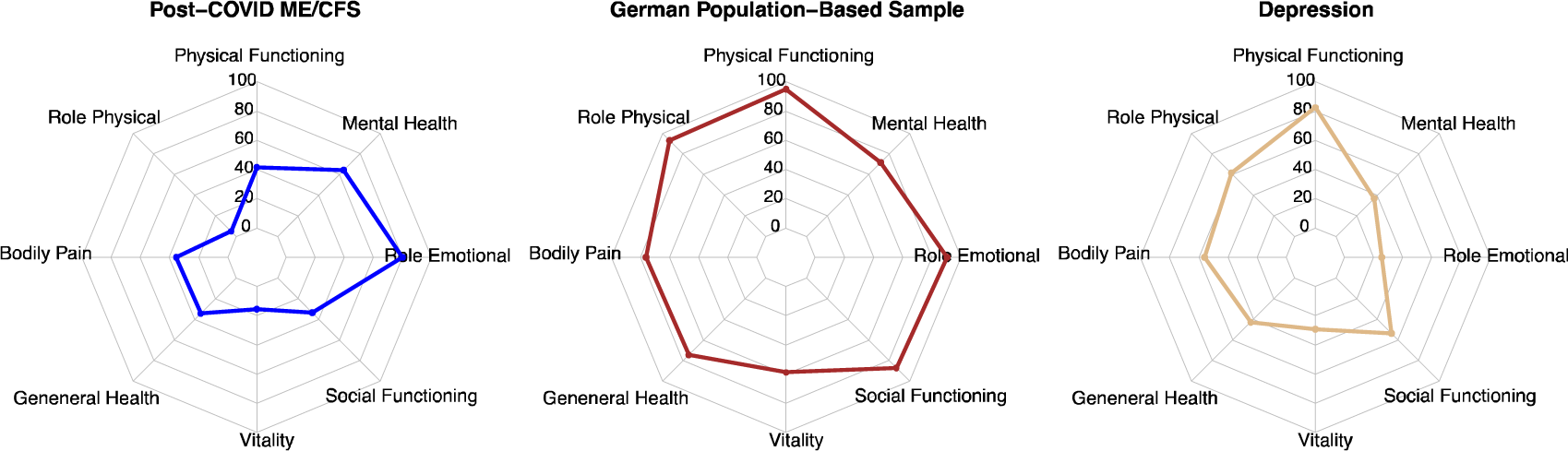
Results from the Short Form 36 Health Survey (SF-36) Spider diagrams display the different dimensions of the Short Form 36 Health Survey (SF-36) for the ten MCFC patients with ME/CFS following COVID-19 (Post-COVID-ME/CFS), the German norm population (age 14 – 20 years) from 1998 [70], and patients with moderate to severe depression (n = 60, mean age 17.5±1.6 years) [72].

**Figure 2a.**
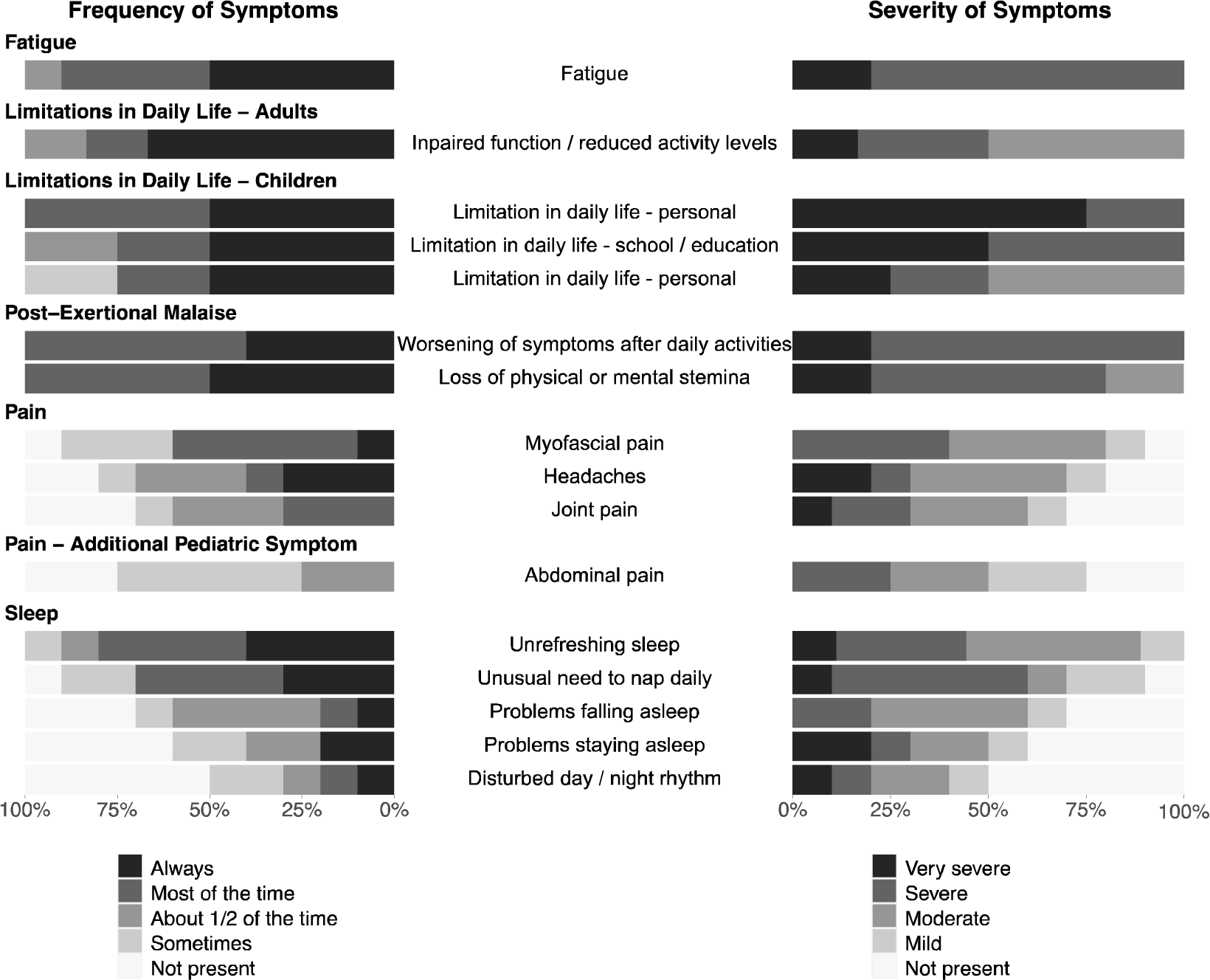
Frequency and Severity of Symptoms. Stacked bar charts represent the frequency and severity of symptoms as indicated on the first page of the Munich Berlin Symptom Questionnaires (MBSQs). Symptoms that are assessed differently in pediatric (n = 4) and adult patients (n = 6) are presented separately, as indicated.

**Figure 2b.**
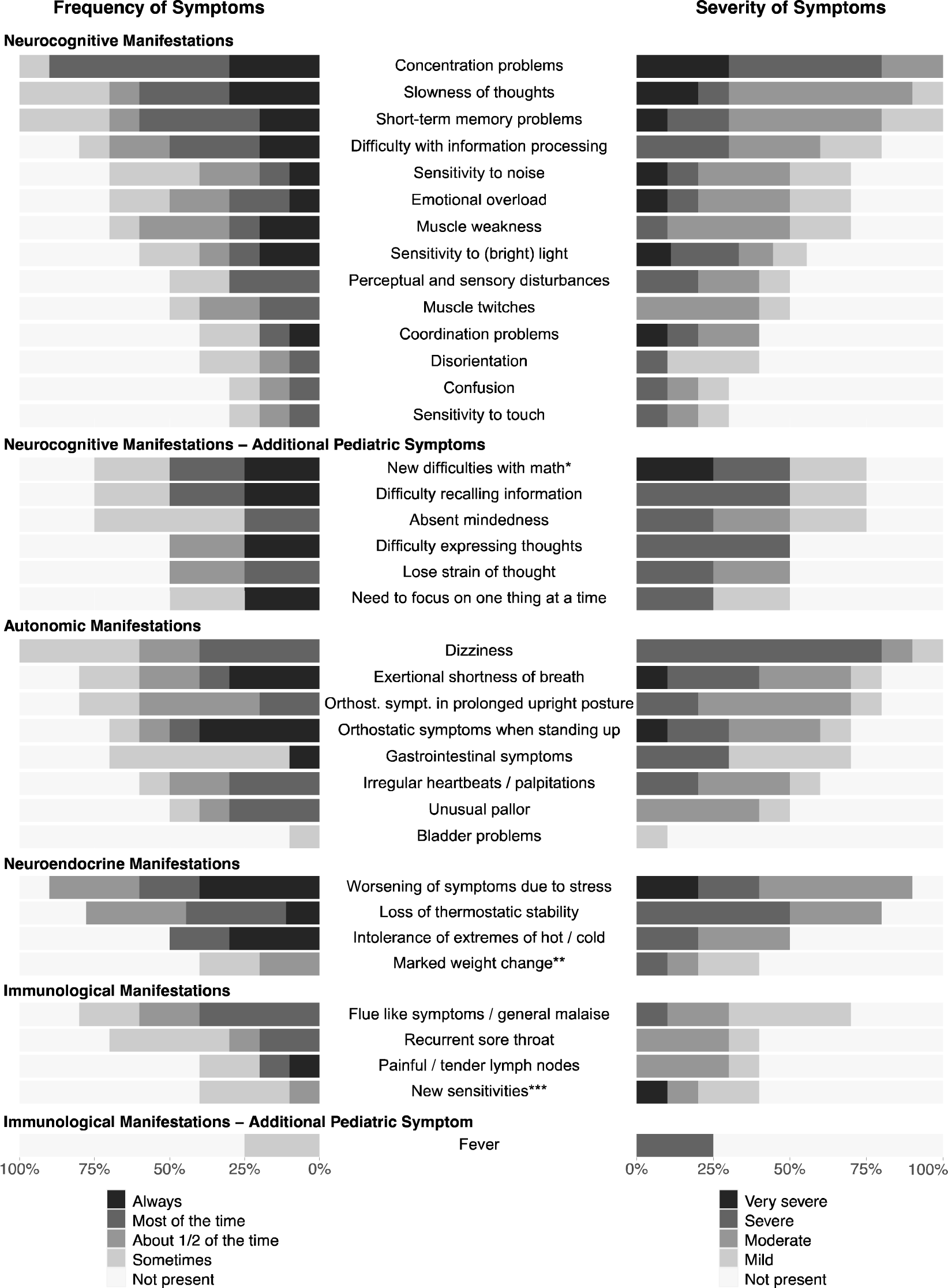
Frequency and Severity of Symptoms. Stacked bar charts display the frequency and severity of symptoms from the second page of the Munich Berlin Symptom Questionnaires (MBSQs). Symptoms that are assessed differently in pediatric (n = 4) and adult patients (n = 6) are presented separately, as indicated. *New difficulties with math or other educational subject; **Marked weight change and / or loss of appetite and / or abnormal appetite; ***New sensitivities to food, medication or chemicals.

The MBSQs were provided to MCFC patients who sought care due to chronic fatigue and PEM. The first ten patients diagnosed with PCC and ME/CFS using the MBSQ included four children and adolescents between 11 and 15 years, and six young adults aged 18 to 25 years, with a male-to-female ratio of 3:7. When presenting at the MCFC, these patients were suffering from PCC symptoms for a period of four to 16 months (**Table 2**).

**Table 2.**
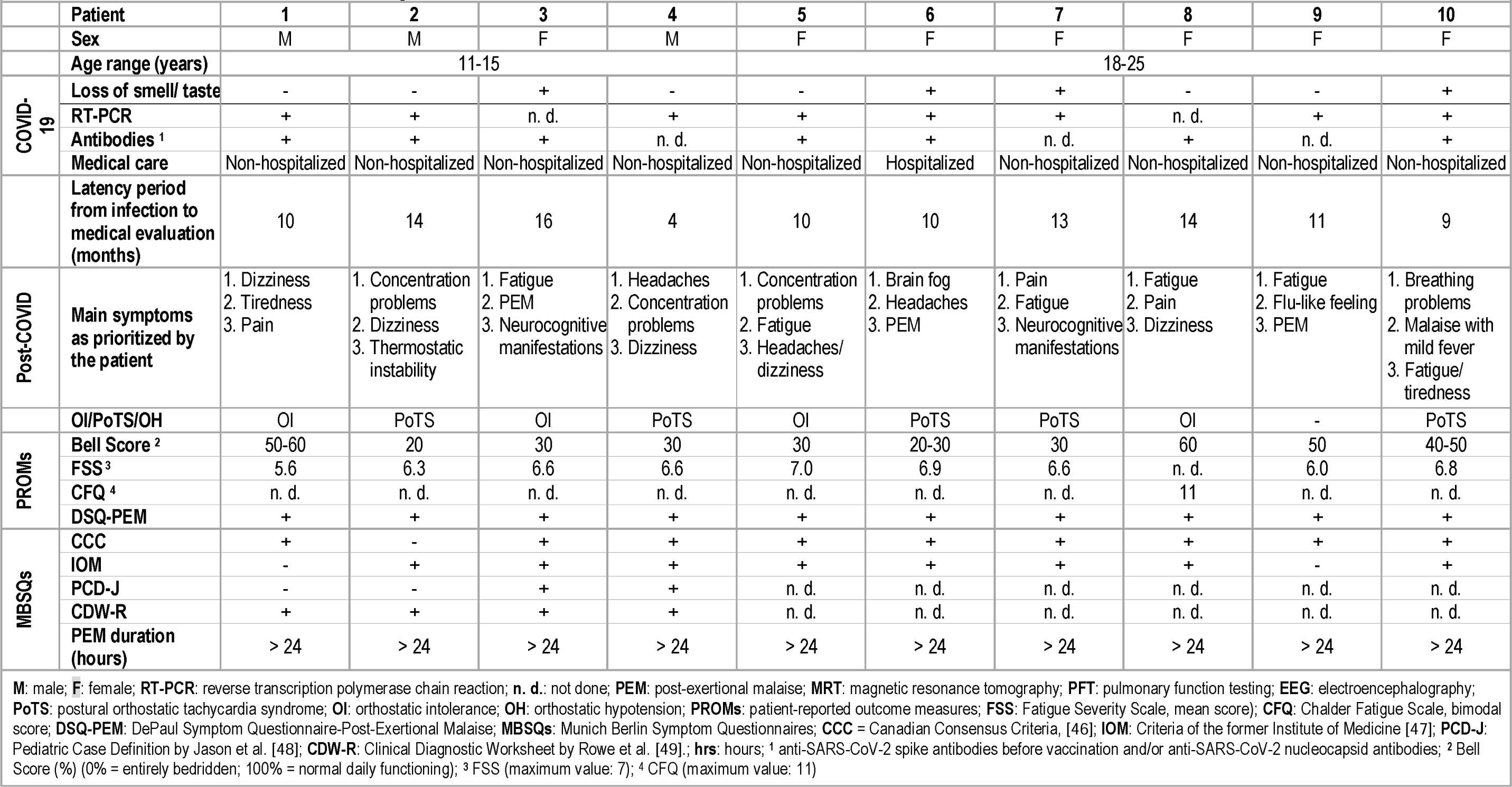
Clinical Data of Patients with MECFS following SARS-CoV-2 infection.

|  |  |  |  |  |  |  |  |  |  |  |  |
| --- | --- | --- | --- | --- | --- | --- | --- | --- | --- | --- | --- |
|  | Patient | 1 | 2 | 3 | 4 | 5 | 6 | 7 | 8 | 9 | 10 |
|  | Sex | M | M | F | M | F | F | F | F | F | F |
|  | Age range (years) | 11-15 |  |  |  | 18-25 |  |  |  |  |  |
| COVID-19 | Loss of smell/ taste | - | - | + | - | - | + | + | - | - | + |
|  | RT-PCR | + | + | n. d. | + | + | + | + | n. d. | + | + |
|  | Antibodies <sup>1</sup> | + | + | + | n. d. | + | + | n. d. | + | n. d. | + |
|  | Medical care | Non-hospitalized | Non-hospitalized | Non-hospitalized | Non-hospitalized | Non-hospitalized | Hospitalized | Non-hospitalized | Non-hospitalized | Non-hospitalized | Non-hospitalized |
|  | Latency period from infection to medical evaluation (months) | 10 | 14 | 16 | 4 | 10 | 10 | 13 | 14 | 11 | 9 |
| Post-COVID | Main symptoms as prioritized by the patient | 1. Dizziness<br>2. Tiredness<br>3. Pain | 1. Concentration problems<br>2. Dizziness<br>3. Thermostatic instability | 1. Fatigue<br>2. PEM<br>3. Neurocognitive manifestations | 1. Headaches<br>2. Concentration problems<br>3. Dizziness | 1. Concentration problems<br>2. Fatigue<br>3. Headaches/dizziness | 1. Brain fog<br>2. Headaches<br>3. PEM | 1. Pain<br>2. Fatigue<br>3. Neurocognitive manifestations | 1. Fatigue<br>2. Pain<br>3. Dizziness | 1. Fatigue<br>2. Flu-like feeling<br>3. PEM | 1. Breathing problems<br>2. Malaise with mild fever<br>3. Fatigue/tiredness |
|  | OI/POTS/OH | OI | POTS | OI | POTS | OI | POTS | POTS | OI | - | POTS |
| PROMs | Bell Score <sup>2</sup> | 50-60 | 20 | 30 | 30 | 30 | 20-30 | 30 | 60 | 50 | 40-50 |
|  | FSS <sup>3</sup> | 5.6 | 6.3 | 6.6 | 6.6 | 7.0 | 6.9 | 6.6 | n. d. | 6.0 | 6.8 |
|  | CFQ <sup>4</sup> | n. d. | n. d. | n. d. | n. d. | n. d. | n. d. | n. d. | 11 | n. d. | n. d. |
|  | DSQ-PEM | + | + | + | + | + | + | + | + | + | + |
| MBSQs | CCC | + | - | + | + | + | + | + | + | + | + |
|  | IOM | - | + | + | + | + | + | + | + | - | + |
|  | PCD-J | - | - | + | + | n. d. | n. d. | n. d. | n. d. | n. d. | n. d. |
|  | CDW-R | + | + | + | + | n. d. | n. d. | n. d. | n. d. | n. d. | n. d. |
|  | PEM duration (hours) | > 24 | > 24 | > 24 | > 24 | > 24 | > 24 | > 24 | > 24 | > 24 | > 24 |

**M:** male; **F:** female; **RT-PCR:** reverse transcription polymerase chain reaction; **n. d.:** not done; **PEM:** post-exertional malaise; **MRT:** magnetic resonance tomography; **PFT:** pulmonary function testing; **EEG:** electroencephalography; **POTS:** postural orthostatic tachycardia syndrome; **OI:** orthostatic intolerance; **OH:** orthostatic hypotension; **PROMs:** patient-reported outcome measures; **FSS:** Fatigue Severity Scale, mean score); **CFQ:** Chalder Fatigue Scale, bimodal score; **DSQ-PEM:** DePaul Symptom Questionnaire-Post-Exertional Malaise; **MBSQs:** Munich Berlin Symptom Questionnaires; **CCC** = Canadian Consensus Criteria, [46]; **IOM:** Criteria of the former Institute of Medicine [47]; **PCD-J:** Pediatric Case Definition by Jason et al. [48]; **CDW-R:** Clinical Diagnostic Worksheet by Rowe et al. [49]; **hrs:** hours; <sup>1</sup> anti-SARS-CoV-2 spike antibodies before vaccination and/or anti-SARS-CoV-2 nucleocapsid antibodies; <sup>2</sup> Bell Score (%) (0% = entirely bedridden; 100% = normal daily functioning); <sup>3</sup> FSS (maximum value: 7); <sup>4</sup> CFQ (maximum value: 11)

9/10 patients were diagnosed with confirmed or probable COVID-19 and 1/10 with asymptomatic SARS-CoV-2 infection between March 2020 and January 2022. 8/10 patients provided a positive SARS-CoV-2 RT-PCR result, and 2/10 patients showed SARS-CoV-2 IgG antibodies without prior COVID-19 vaccination, together with a history of COVID-19-like symptoms. 9/10 patients were initially treated as outpatients, while one adult was hospitalized for four days due to a pre-syncopal episode in the context of COVID-19. An initial loss of smell and/or taste was reported by one adolescent and three adults (**Table 2**).

Pre-existing medical conditions were present in almost all (9/10) patients, including bronchial asthma (4/10), hypothyroidism (2/10), Grave’s disease with hyperthyroidism (1/10), allergies (2/10), attention deficit disorder (1/10), migraine with aura (1/10), history of meningitis (1/10), or Alport’s syndrome (1/10) (**Table 2**).

The comprehensive diagnostic workup did not reveal any other explanation for the reported complex and debilitating symptoms. Physical and neurological examination, as well as ECG and UCG, showed normal findings in all patients. A cardiac MRI performed on an adult patient revealed a condition after perimyocarditis. In two patients, the PFT showed signs of a hyper-responsive bronchial system, one of which reported pre-existing asthma bronchiale. A cranial MRI was indicated by neurologists in nine and an EEG in eight patients, with normal results except a stable, benign, cystic CNS lesion in one and a transient theta wave slowing in another patient. None of these findings were considered to explain the complex symptoms. 9/10 patients complained of OI, with 5/10 patients meeting the diagnostic criteria for PoTS.

All patients reported significant fatigue in the FSS (9/9) or CFQ (1/1) and screened positive for PEM in the DSQ-PEM, with a PEM duration of more than 24 hours (**Table 2**). Daily functioning measured by the Bell Score ranged between 20% to 60% (median: 30, IQR: 30 - 48.75). All patients considered their symptoms as very debilitating and had significant difficulties with schooling, apprenticeship, or academic studies.

Results from the SF-36 displayed impairment in all dimensions (**Figure 1**) relative to the German norm sample for the age of 14 to 20 years [70]. The physical component summary (PCS) was markedly reduced in our group of ME/CFS patients compared to the German norm population, with a mean score of 24.9 compared to 53.4 (P < 0.001). The mean mental component summary score was 44.9 versus 45.0, respectively (P = 0.982) [70].

All patients experienced substantial reductions in occupational, educational, and/or personal activities, indicated by scoring at or below at least two of the three following subscale cut-offs on the SF-36: role physical ≤ 50, social functioning ≤ 62.5, and vitality ≤ 35, as required by the original CCC and the PCD-J [71]. Compared to patients suffering from mild to moderate depression (n = 60, mean age 17.5 ±1.6 years) [72], our ME/CFS patients had significantly reduced scores in the subscales physical functioning (P < 0.001), role physical (P < 0.001), bodily pain (P < 0.001), vitality (P = 0.002), and social functioning (P = 0.016). Only in the two subscales role emotional (P < 0.001) and mental health (P < 0.001) our patients scored significantly higher than the control group of adolescents and young adults with moderate to severe depression. The subscale general health was not significantly different (P = 0.082) (**Figure 1**).

All patients fulfilled at least one ME/CFS case definition addressed in the MBSQ. One child fulfilled the CCC and the PDW-R but not the IOM and the PCD-J. Two adolescents met all four sets of criteria, while one met only the broader PDW-R and IOM criteria. All adults fulfilled the CCC, but one did not match the IOM criteria since sleep was not recognized as “unrefreshing” (**Table 2**).

The most common ME/CFS symptoms were fatigue (10/10), limitations in daily life (10/10), long-lasting PEM (10/10), unrefreshing sleep (9/10), neurocognitive manifestations (10/10) (e.g., concentration and memory problems), and dizziness (6/10). The most bothering symptoms, the total amount of symptoms, as well as the frequency and severity of symptoms varied individually (**Table 2**, **Figure 2**).

## DISCUSSION

The MBSQs and SSSs are novel, age-adapted, concise diagnostic tools developed to facilitate the evaluation of ME/CFS criteria in patients with fatigue following COVID-19 and beyond. We reported ten young PCC patients who were diagnosed with ME/CFS using the MBSQ. To our knowledge, this is the first report on ME/CFS in people with PCC younger than 18 years.

Very little is known about the prevalence of severe PCC in children and adolescents. A survey over a four-week period ending 30 March 2023 on the prevalence of ongoing symptoms following SARS-CoV-2 infection in U.K. households indicated that sequelae defined as “limiting day-to-day activities” manifested less often in children aged 2-11 years (0.1%) compared to adolescents and young adults (12-24 years) (0.26 - 0.33%) and adults aged ≥ 25 years (0.46 - 0.92%) [73]. However, early in the pandemic, a report from Sweden indicated that pediatric PCC can compromise school education for several months [74], and a single 19-year-old adolescent with post-COVID-ME/CFS was documented in the US [27; 28]. Meanwhile, additional pediatric patients were diagnosed with ME/CFS at our MCFC and at several partner sites of our recently implemented multicenter long COVID (NCT05638724) and ME/CFS registries (NCT05778006), as will be reported in more detail (unpublished results).

ME/CFS, in general, is well documented in children and adolescents [49]. In a pre-pandemic pediatric cohort from Australia, ME/CFS was reported to have followed infections in up to 80% of cases, with EBV infection accounting for 40% of cases [32]. 12.9%, 7.3%, and 4.3% of adolescents in a pre-pandemic U.S. cohort presented with ME/CFS as defined by the PCD-J criteria at six, 12, and 24 months after EBV-induced infectious mononucleosis [75]. ME/CFS defined by meeting at least one of three case definitions (Fukuda, IOM, CCC) manifested in 23% of U.S. college students following symptomatic primary EBV infection, with 8% of the cohort fulfilling the CCC [76]. The prognosis of ME/CFS was reported to be more favorable in children and adolescents compared to adults, with recovery rates of 38% after five years and 68% after ten years and a mean illness duration of five (range 1–15) years in those who recovered [32]. Post-viral ME/CFS and partial recovery thereof was also documented at the MCFC [77]. Taken together, pediatric ME/CFS following infection with SARS-CoV-2 was not unexpected and might be transient, at least in some cases, if diagnosed and treated in a correct and timely way.

Reports of ME/CFS after SARS-CoV-2 infection in adults have been published, including reports from Germany [21; 25; 27]. However, little is known about the pediatric population. To our knowledge, this is the first report on ME/CFS in PCC patients aged less than 18 years, including children as young as 11 years. All reported patients had developed long-lasting typical symptoms after asymptomatic SARS-CoV-2 infection or mild to moderate courses of documented or probable COVID-19 with no other medical explanation and impairment of daily life, according to the WHO definition of PCC [11]. A substantial reduction in occupational, educational, and/or personal activities of these patients was confirmed by a Bell score of ≤ 60% and by scoring at or below at least two of three subscale cut-offs on the SF-36 (role physical ≤ 50, social functioning ≤ 62.5, and vitality ≤ 35) [71].

In line with current evidence indicating a lower prevalence of severe PCC in children younger than 12 years compared to adolescents and adults [73], 9 of 10 of our patients were older than 12 years. At the MCFC, we are regularly seeing young adults up to the age of 20 years since their healthcare needs, in general, do not substantially differ from those of older adolescents and include features such as school or peer group integration that might not be fully covered by healthcare institutions for adult patients. Moreover, patients turning 18 years had been excluded from other pediatric PCC follow-up studies indicating a possible lack of data in this age group [78].

The general lack of data regarding ME/CFS following SARS-CoV-2 and other infections may, in part, be explained by insufficient disease-specific knowledge and experience [49; 54]. Further, the comparison of published data is challenging due to the different ME/CFS case definitions used worldwide [79; 80]. Thus, harmonization of diagnostic criteria for ME/CFS is urgently needed. High time and cost expenses for the diagnostic workup, moreover, may prevent clinicians from diagnosing ME/CFS and as a result these patients often get no adequate care.

The MBSQs and SSSs were developed as concise tools that can facilitate and harmonize diagnostic workup in clinical and research centers currently faced with PCC and other post-infection syndromes. They provide a standardized basis for a semi-structured medical interview. The interview guarantees to rule out any misunderstanding, especially concerning PEM, the key feature of ME/CFS. Thus, the MBSQs may be pre-filled by patients at home but should not be used as PROMs.

Our approach was based on the DSQs developed as PROMs by L.A. Jason and colleagues to evaluate ME/CFS diagnosis and associated features in studies with adults, adolescents, and children [57]. Like the DSQs, the MBSQs offer Likert scales for the quantification of symptoms, with a threshold of ≥ 2 for both frequency and severity to indicate diagnostic relevance. Moreover, as introduced by the DSQs, the SSSs provide an algorithm to evaluate different case definitions using a single questionnaire.

In contrast to the DSQs, the MBSQs neither address demographics nor do they evaluate the medical, occupational, and social history since many PCC centers might want to follow their own respective questionnaires and since some items of the DSQs are related to specific settings in the US. Furthermore, in contrast to the DSQ-2, the MBSQs do not address the comprehensive international consensus criteria (ME-ICC) because they were not recommended by the European Network for ME/CFS (EUROMENE) [31].

Importantly, the MBSQs are setting a cut-off at ≥14 hours regarding the PEM duration for the CCC as well as for the PCD-J and CDW-R criteria while leaving the IOM criteria without. Previous ME/CFS studies indicated that PEM lasts longer than 24 hours in most patients [24]. Some ME/CFS case definitions required a duration of at least 24 hours, including the CDW-R (“Recovery takes more than 24h”) [49]. The original publications of the CCC and the PCD-J stated that “there is a pathologically slow recovery period–usually 24 hours or longer” (CCC) [46] and that “the recovery is slow, often taking 24 hours or longer” (PCD-J) [48]), respectively. For the MBSQs, we chose a PEM duration cut-off at ≥ 14 hours since it included more ME/CFS patients than a cut-off at ≥ 24 hours but still excluded the majority of patients with other chronic diseases in a previous study [24]. However, since the MBSQs offer various durations of PEM, they allow for the investigation of subgroups for research purposes.

The broader IOM criteria included in the MBSQ were recommended for clinical diagnosis by the EUROMENE and by the CDC in the US. The IOM criteria lack, however, a definition of the length of PEM and do not require cognitive symptoms. In PCS, the assessment of the length of PEM is, however, important as it defines subgroups of patients with different biomarker profiles and clinical courses [21; 81-84]. Uniform use of one case definition in clinical practice would allow for reasonable comparability of healthcare data on ME/CFS worldwide, including ME/CFS in the context of PCC.

The MBSQs have several limitations. First, results have not been compared with results from other questionnaires investigating ME/CFS case definitions such as e.g. the DSQ-2 for reasons of practicability. Patients at the MCFC have to fill in a long list of questionnaires which is challenging many of them to an extent that prevents adding additional questionnaires. However, we are recommending the MBSQs not as a PROM but as a tool to facilitate a structured medical interview which can be recognized as the gold standard for evaluating diagnostic ME/CFS criteria. A comparison of results from pre-filled MBSQs, with MBSQ results from the medical visits as well as with results from other questionnaires, will be an important future goal. Second, the low number and heterogeneity of cases regarding age, gender, and pre-existing morbidity did not allow for correlation and consistency analyses of MBSQ results. This will be aimed at using results from a larger group of future patients. Third and most importantly, a structured medical interview based on well-designed questionnaires cannot substitute for a future diagnostic biomarker for ME/CFS and/or PEM. Without a diagnostic biomarker, diagnosing the complex symptom PEM and PEM duration will remain challenging, especially in patients who are largely preventing long-lasting “crashes” by successful pacing. Last but not least, here we only report on patients who eventually met any ME/CFS case definition. In ongoing studies, we are evaluating the MBSQ in the context of healthy individuals and patients with other chronic diseases.

Taken together, the MBSQs and SSSs were developed to standardize and accelerate ME/CFS diagnosis at any age in clinical practice and research and were successfully applied to children, adolescents, and young adults with PCC. Standardization in PCC and ME/CFS research is urgently needed to compare clinical studies, identify biomarkers, and eventually select and develop specific treatment approaches [85].

## Conclusion

We have developed and successfully applied a set of novel diagnostic questionnaire and scoring sheets to identify children, adolescents, and adults with ME/CFS following SARS-CoV-2 infection and beyond. These questionnaires can aid clinicians in assessing up to four case definitions of ME/CFS in a quantitative and standardized manner. The questionnaires include the broader IOM criteria recommended by experts in the U.S. and Europe for clinical care as well as the stricter CCC and, for children and adolescents, two additional pediatric case definitions. These novel tools allow for assessing the frequency and severity of ME/CFS symptoms as well as the duration of PEM and thereby support further research on these features in the context of ME/CFS diagnosis. In sum, we expect the MBSQs to facilitate patient care and research in the context of ME/CFS in pediatrics and beyond.

## Supporting information

MBSQs and Supplementary Scoring Sheets

## Abbreviations

ANA: antinuclear antibodies
CCC: Canadian consensus criteria
CDC: Centers of Disease Control and Prevention
CDW-R: clinical diagnostic worksheet of P.C. Rowe et al. (2017)
CFC: Charité Fatigue Center
COVID-19: coronavirus disease 2019
CRP: C-reactive protein
DSQ: DePaul symptom questionnaire
EBV: Epstein-Barr virus
ECG: electrocardiography
EEG: electroencephalography
EUROMENE: European Network on ME/CFS
GET: graded exercise therapy
HR: heart rate
HRQoL: health-related quality of life
IOM: Institute of Medicine
KBV: German National Association of Statutory Health Insurance Physicians
MBSQ: Munich Berlin Symptom Questionnaire
MCFC: MRI Chronic Fatigue Center for Young People
ME/CFS: myalgic encephalomyelitis / chronic fatigue syndrome
MRI: TUM university hospital (Klinikum rechts der Isar)
MRT: magnetic resonance tomography
NICE: National Institute for Health and Care Excellence
OH: orthosthatic hypotension
OI: orthosthatic intolerance
PASC: post-acute sequelae of COVID-19
PCC: post-COVID-19 condition
PCD-J: pediatric case definition by L.A. Jason et al. (2006)
PEM: post-exertional malaise
PFT: pulmonary function testing
PoTS: postural tachcardia syndrome
PROM: patient-reported outcome measure
RT-PCR: reverse transcriptase - polymerase chain reaction
SARS-CoV-2: severe acute respiratory coronavirus type 2
SEID: systemic exertion intolerance disease criteria
SF-36: Short Form 36
SSS: Supplementary Scoring Sheet
UCG: ultrasound cardiography
WHO: World Health Organisation

## Acknowledgments

We thank all patients who participated in the project as well as their families, for supporting the participation.

## STATEMENTS AND DECLARATIONS

### Author contribution statement

Conceptualization: L.C.P, K.W., R.P., K.G., J.P., A.L., U.B.. Methodology: L.C.P, K.W., R.P., K.G., J.P., A.L., C.S., D.H., L.F., U.B.. Data curation, L.C.P., R.P., A.V.. Formal analysis: L.C.P., R.P., L.M. Draft writing: L.C.P, K.W., R.P.. Editing: L.C.P, R.P., M.H., M.A., A.V., A.L., S.S., S.A., C.W., D.H., L.F., L.M., C.S., U.B.. Figures: R.P., L.M.. Supervision, U.B.. Acquisition of Funding: U.B.. All authors contributed to the final manuscript. All authors have read and agreed to the published version of the manuscript.

### Ethical Approval

The study was conducted according to the guidelines of the Declaration of Helsinki and approved by the Ethics Committee of the University Hospital of the Technical University of Munich (116/21, 511/21).

### Consent to Participate

Written informed consent was obtained from all subjects (or their legal guardian) involved in the study.

### Data Availability Statement

The data that support the findings of this study are available from the corresponding author, U.B., upon reasonable request.

## REFERENCES

1. Pei S, Yamana TK, Kandula S, Galanti M, Shaman J (2021) Burden and characteristics of COVID-19 in the United States during 2020. Nature 598:338–341

2. Msemburi W, Karlinsky A, Knutson V, Aleshin-Guendel S, Chatterji S, Wakefield J (2023) The WHO estimates of excess mortality associated with the COVID-19 pandemic. Nature 613:130–137

3. Mueller MR, Ganesh R, Hurt RT, Beckman TJ (2023) Post-COVID Conditions. Mayo Clin Proc 98:1071–1078

4. Davis HE, McCorkell L, Vogel JM, Topol EJ (2023) Long COVID: major findings, mechanisms and recommendations. Nat Rev Microbiol 21:133–146

5. Nalbandian A, Sehgal K, Gupta A, Madhavan MV, McGroder C, Stevens JS, Cook JR, et al. (2021) Post-acute COVID-19 syndrome. Nat Med 27:601–615

6. Carfì A, Bernabei R, Landi F (2020) Persistent Symptoms in Patients After Acute COVID-19. Jama 324:603–605

7. Han Q, Zheng B, Daines L, Sheikh A (2022) Long-Term Sequelae of COVID-19: A Systematic Review and Meta-Analysis of One-Year Follow-Up Studies on Post-COVID Symptoms. Pathogens 11

8. Zimmermann P, Pittet LF, Curtis N (2022) The Challenge of Studying Long COVID: An Updated Review. Pediatr Infect Dis J 41:424–426

9. Zimmermann P, Pittet LF, Curtis N (2021) How Common is Long COVID in Children and Adolescents? Pediatr Infect Dis J 40:e482–e487

10. Office for National Statistics (2023) Prevalence of ongoing symptoms following coronavirus (COVID-19) infection in the UK: 2 February 2023.

11. World Health Organization (2023) A clinical case definition for post COVID-19 condition in children and adolescents by expert consensus. World Health Organization

12. World Health Organization (2021) A clinical case definition of post COVID-19 condition by a Delphi consensus. World Health Organization, Geneva, Switzerland

13. Scharf RE, Anaya JM (2023) Post-COVID Syndrome in Adults-An Overview. Viruses 15

14. Pellegrino R, Chiappini E, Licari A, Galli L, Marseglia GL (2022) Prevalence and clinical presentation of long COVID in children: a systematic review. Eur J Pediatr 181:3995–4009

15. Molteni E, Sudre CH, Canas LS, Bhopal SS, Hughes RC, Antonelli M, Murray B, Kläser K, Kerfoot E, Chen L, Deng J, Hu C, Selvachandran S, Read K, Capdevila Pujol J, Hammers A, Spector TD, Ourselin S, Steves CJ, Modat M, Absoud M, Duncan EL (2021) Illness duration and symptom profile in symptomatic UK school-aged children tested for SARS-CoV-2. Lancet Child Adolesc Health 5:708–718

16. Stephenson T, Pinto Pereira SM, Shafran R, de Stavola BL, Rojas N, McOwat K, Simmons R, Zavala M, O’Mahoney L, Chalder T, Crawley E, Ford TJ, Harnden A, Heyman I, Swann O, Whittaker E, Consortium C, Ladhani SN (2022) Physical and mental health 3 months after SARS-CoV-2 infection (long COVID) among adolescents in England (CLoCk): a national matched cohort study. Lancet Child Adolesc Health 6:230–239

17. Nittas V, Gao M, West EA, Ballouz T, Menges D, Wulf Hanson S, Puhan MA (2022) Long COVID Through a Public Health Lens: An Umbrella Review. Public Health Rev 43:1604501

18. Halpin SJ, McIvor C, Whyatt G, Adams A, Harvey O, McLean L, Walshaw C, Kemp S, Corrado J, Singh R, Collins T, O’Connor RJ, Sivan M (2021) Postdischarge symptoms and rehabilitation needs in survivors of COVID-19 infection: A cross-sectional evaluation. J Med Virol 93:1013–1022

19. Townsend L, Dyer AH, Jones K, Dunne J, Mooney A, Gaffney F, O’Connor L, Leavy D, O’Brien K, Dowds J, Sugrue JA, Hopkins D, Martin-Loeches I, Ni Cheallaigh C, Nadarajan P, McLaughlin AM, Bourke NM, Bergin C, O’Farrelly C, Bannan C, Conlon N (2020) Persistent fatigue following SARS-CoV-2 infection is common and independent of severity of initial infection. PLoS One 15:e0240784

20. Ceban F, Ling S, Lui LMW, Lee Y, Gill H, Teopiz KM, Rodrigues NB, Subramaniapillai M, Di Vincenzo JD, Cao B, Lin K, Mansur RB, Ho RC, Rosenblat JD, Miskowiak KW, Vinberg M, Maletic V, McIntyre RS (2022) Fatigue and cognitive impairment in Post-COVID-19 Syndrome: A systematic review and meta-analysis. Brain Behav Immun 101:93–135

21. Kedor C, Freitag H, Meyer-Arndt L, Wittke K, Hanitsch LG, Zoller T, Steinbeis F, Haffke M, Rudolf G, Heidecker B, Bobbert T, Spranger J, Volk H-D, Skurk C, Konietschke F, Paul F, Behrends U, Bellmann-Strobl J, Scheibenbogen C (2022) A prospective observational study of post-COVID-19 chronic fatigue syndrome following the first pandemic wave in Germany and biomarkers associated with symptom severity. Nature Communications 13:5104

22. Jason LA, Dorri JA (2022) ME/CFS and Post-Exertional Malaise among Patients with Long COVID. Neurol Int 15:1–11

23. Choutka J, Jansari V, Hornig M, Iwasaki A (2022) Unexplained post-acute infection syndromes. Nature Medicine 28:911–923

24. Cotler J, Holtzman C, Dudun C, Jason LA (2018) A Brief Questionnaire to Assess Post-Exertional Malaise. Diagnostics (Basel) 8

25. Bonilla H, Quach TC, Tiwari A, Bonilla AE, Miglis M, Yang PC, Eggert LE, Sharifi H, Horomanski A, Subramanian A, Smirnoff L, Simpson N, Halawi H, Sum-ping O, Kalinowski A, Patel ZM, Shafer RW, Geng LN (2023) Myalgic Encephalomyelitis/Chronic Fatigue Syndrome is common in post-acute sequelae of SARS-CoV-2 infection (PASC): Results from a post-COVID-19 multidisciplinary clinic. Front Neurol 14:1090747

26. González-Hermosillo JA, Martínez-López JP, Carrillo-Lampón SA, Ruiz-Ojeda D, Herrera-Ramírez S, Amezcua-Guerra LM, Martínez-Alvarado MDR (2021) Post-Acute COVID-19 Symptoms, a Potential Link with Myalgic Encephalomyelitis/Chronic Fatigue Syndrome: A 6-Month Survey in a Mexican Cohort. Brain Sci 11

27. Petracek LS, Suskauer SJ, Vickers RF, Patel NR, Violand RL, Swope RL, Rowe PC (2021) Adolescent and Young Adult ME/CFS After Confirmed or Probable COVID-19. Front Med (Lausanne) 8:668944

28. Petracek LS, Broussard CA, Swope RL, Rowe PC (2023) A Case Study of Successful Application of the Principles of ME/CFS Care to an Individual with Long COVID. Healthcare (Basel) 11

29. Jason LA, Johnson M, Torres C (2023) Pediatric Post-Acute Sequelae of SARS-CoV-2 infection. Fatigue: Biomedicine, Health & Behavior:1–11

30. Rasa S, Nora-Krukle Z, Henning N, Eliassen E, Shikova E, Harrer T, Scheibenbogen C, Murovska M, Prusty BK (2018) Chronic viral infections in myalgic encephalomyelitis/chronic fatigue syndrome (ME/CFS). J Transl Med 16:268

31. Nacul L, Authier FJ, Scheibenbogen C, Lorusso L, Helland IB, Martin JA, Sirbu CA, et al. (2021) European Network on Myalgic Encephalomyelitis/Chronic Fatigue Syndrome (EUROMENE): Expert Consensus on the Diagnosis, Service Provision, and Care of People with ME/CFS in Europe. Medicina (Kaunas) 57

32. Rowe KS (2019) Long Term Follow up of Young People With Chronic Fatigue Syndrome Attending a Pediatric Outpatient Service. Front Pediatr 7:21

33. Estévez-López F, Mudie K, Wang-Steverding X, Bakken IJ, Ivanovs A, Castro-Marrero J, Nacul L, Alegre J, Zalewski P, Słomko J, Strand EB, Pheby D, Shikova E, Lorusso L, Capelli E, Sekulic S, Scheibenbogen C, Sepúlveda N, Murovska M, Lacerda E (2020) Systematic Review of the Epidemiological Burden of Myalgic Encephalomyelitis/Chronic Fatigue Syndrome Across Europe: Current Evidence and EUROMENE Research Recommendations for Epidemiology. J Clin Med 9

34. Komaroff AL, Lipkin WI (2021) Insights from myalgic encephalomyelitis/chronic fatigue syndrome may help unravel the pathogenesis of postacute COVID-19 syndrome. Trends Mol Med 27:895–906

35. Behnood SA, Shafran R, Bennett SD, Zhang AXD, O’Mahoney LL, Stephenson TJ, Ladhani SN, De Stavola BL, Viner RM, Swann OV (2022) Persistent symptoms following SARS-CoV-2 infection amongst children and young people: A meta-analysis of controlled and uncontrolled studies. J Infect 84:158–170

36. Tsampasian V, Elghazaly H, Chattopadhyay R, Debski M, Naing TKP, Garg P, Clark A, Ntatsaki E, Vassiliou VS (2023) Risk Factors Associated With Post-COVID-19 Condition: A Systematic Review and Meta-analysis. JAMA Intern Med 183:566–580

37. Nacul LC, Lacerda EM, Pheby D, Campion P, Molokhia M, Fayyaz S, Leite JC, Poland F, Howe A, Drachler ML (2011) Prevalence of myalgic encephalomyelitis/chronic fatigue syndrome (ME/CFS) in three regions of England: a repeated cross-sectional study in primary care. BMC Med 9:91

38. Jason LA, Richman JA, Rademaker AW, Jordan KM, Plioplys AV, Taylor RR, McCready W, Huang CF, Plioplys S (1999) A community-based study of chronic fatigue syndrome. Arch Intern Med 159:2129–2137

39. Jason L, Mirin A (2021) Updating the National Academy of Medicine ME/CFS prevalence and economic impact figures to account for population growth and inflation. Fatigue: Biomedicine, Health & Behavior 9:9–13

40. Lim E-J, Ahn Y-C, Jang E-S, Lee S-W, Lee S-H, Son C-G (2020) Systematic review and meta-analysis of the prevalence of chronic fatigue syndrome/myalgic encephalomyelitis (CFS/ME). Journal of translational medicine 18:1–15

41. Crawley EM, Emond AM, Sterne JAC (2011) Unidentified Chronic Fatigue Syndrome/myalgic encephalomyelitis (CFS/ME) is a major cause of school absence: surveillance outcomes from school-based clinics. BMJ Open 1:e000252

42. Jason LA, Katz BZ, Sunnquist M, Torres C, Cotler J, Bhatia S (2020) The Prevalence of Pediatric Myalgic Encephalomyelitis/Chronic Fatigue Syndrome in a Communityl.ZBased Sample. Child Youth Care Forum 49:563–579

43. Mirin AA, Dimmock ME, Jason LA (2022) Updated ME/CFS prevalence estimates reflecting post-COVID increases and associated economic costs and funding implications. Fatigue: Biomedicine, Health & Behavior 10:83–93

44. Roessler M, Tesch F, Batram M, Jacob J, Loser F, Weidinger O, Wende D, et al. (2022) Post-COVID-19-associated morbidity in children, adolescents, and adults: A matched cohort study including more than 157,000 individuals with COVID-19 in Germany. PLoS Med 19:e1004122

45. Kassenärztliche Bundesvereinigung (2023) Öffentliche Anhörung im Ausschuss für Gesundheit des deutschen Bundestages am 19. April 2023. Stellungnahme der KBV zum Antrag der CDU/CSU-Bundestagsfraktion “ME/CFS-Betroffenen sowie deren Angehörigen helfen – Für eine bessere Gesundheits-sowie Therapieversorgung, Aufklärung und Anerkennung.

46. Carruthers BM, Jain AK, De Meirleir KL, Peterson DL, Klimas NG, Lerner AM, Bested AC, Flor-Henry P, Joshi P, Powles ACP, Sherkey JA, van de Sande MI (2003) Myalgic Encephalomyelitis/Chronic Fatigue Syndrome. Journal of Chronic Fatigue Syndrome 11:7–115

47. Clayton EW (2015) Beyond myalgic encephalomyelitis/chronic fatigue syndrome: an IOM report on redefining an illness. Jama 313:1101–1102

48. Jason LA, Jordan K, Miike T, Bell DS, Lapp C, Torres-Harding S, Rowe K, Gurwitt A, De Meirleir K, Van Hoof ELS (2006) A Pediatric Case Definition for Myalgic Encephalomyelitis and Chronic Fatigue Syndrome. Journal of Chronic Fatigue Syndrome 13:1–44

49. Rowe PC, Underhill RA, Friedman KJ, Gurwitt A, Medow MS, Schwartz MS, Speight N, Stewart JM, Vallings R, Rowe KS (2017) Myalgic Encephalomyelitis/Chronic Fatigue Syndrome Diagnosis and Management in Young People: A Primer. Front Pediatr 5:121

50. National Institute for Health and Care Excellence (2021) Myalgic encephalomyelitis (or encephalopathy)/chronic fatigue syndrome: diagnosis and management.

51. Bateman L, Bested AC, Bonilla HF, Chheda BV, Chu L, Curtin JM, Dempsey TT, Dimmock ME, Dowell TG, Felsenstein D, Kaufman DL, Klimas NG, Komaroff AL, Lapp CW, Levine SM, Montoya JG, Natelson BH, Peterson DL, Podell RN, Rey IR, Ruhoy IS, Vera-Nunez MA, Yellman BP (2021) Myalgic Encephalomyelitis/Chronic Fatigue Syndrome: Essentials of Diagnosis and Management. Mayo Clinic Proceedings 96:2861–2878

52. National Institute for Health and Care Excellence: Clinical Guidelines (2021) COVID-19 rapid guideline: managing the long-term effects of COVID-19. National Institute for Health and Care Excellence (UK), London, England

53. Brown MM, Bell DS, Jason LA, Christos C, Bell DE (2012) Understanding long-term outcomes of chronic fatigue syndrome. Journal of clinical psychology 68:1028–1035

54. Hng KN, Geraghty K, Pheby DFH (2021) An Audit of UK Hospital Doctors’ Knowledge and Experience of Myalgic Encephalomyelitis. Medicina (Kaunas) 57

55. Froehlich L, Hattesohl DBR, Jason LA, Scheibenbogen C, Behrends U, Thoma M (2021) Medical Care Situation of People with Myalgic Encephalomyelitis/Chronic Fatigue Syndrome in Germany. Medicina (Kaunas) 57

56. Froehlich L, Niedrich J, Hattesohl DBR, Behrends U, Kedor C, Haas JP, Stingl M, Scheibenbogen C (2023) Evaluation of a webinar to increase health professionals’ knowledge about Myalgic Encephalomyelitis/ Chronic Fatigue Syndrome (ME/CFS). Manuscript submitted for publication

57. Jason LA, Sunnquist M (2018) The Development of the DePaul Symptom Questionnaire: Original, Expanded, Brief, and Pediatric Versions. Front Pediatr 6:330

58. Bedree H, Sunnquist M, Jason LA (2019) The DePaul Symptom Questionnaire-2: A Validation Study. Fatigue 7:166–179

59. Bavarian State Ministry of Health and Care (2021) Modellprojekt „Post-COVID Kids Bavaria“. Teilprojekt 2 „Post-COVID Kids Bavaria - PCFC“ (Post-COVID Fatigue Center). Bavarian State Ministry of Health and Care

60. Krupp LB, LaRocca NG, Muir-Nash J, Steinberg AD (1989) The Fatigue Severity Scale: Application to Patients With Multiple Sclerosis and Systemic Lupus Erythematosus. Archives of Neurology 46:1121–1123

61. Chalder T, Berelowitz G, Pawlikowska T, Watts L, Wessely S, Wright D, Wallace EP (1993) Development of a fatigue scale. J Psychosom Res 37:147–153

62. Bell DS (1994) The doctor’s guide to chronic fatigue syndrome: understanding, treating, and living with CFIDS. Da Capo Press

63. Ware JE, Jr., Sherbourne CD (1992) The MOS 36-item short-form health survey (SF-36). I. Conceptual framework and item selection. Med Care 30:473–483

64. Lerdal A, Wahl A, Rustoen T, Hanestad BR, Moum T (2005) Fatigue in the general population: a translation and test of the psychometric properties of the Norwegian version of the fatigue severity scale. Scand J Public Health 33:123–130

65. Jackson C (2015) The Chalder Fatigue Scale (CFQ 11). Occup Med (Lond) 65:86

66. Lee J, Vernon SD, Jeys P, Ali W, Campos A, Unutmaz D, Yellman B, Bateman L (2020) Hemodynamics during the 10-minute NASA Lean Test: evidence of circulatory decompensation in a subset of ME/CFS patients. J Transl Med 18:314

67. World Health Organization (2018) ICD-11 MMS. International Classification of Diseases for Mortality and Morbidity Statistics. 8D89.2 Postural orthostatic tachycardia syndrome

68. Vernino S, Bourne KM, Stiles LE, Grubb BP, Fedorowski A, Stewart JM, Arnold AC, et al. (2021) Postural orthostatic tachycardia syndrome (POTS): State of the science and clinical care from a 2019 National Institutes of Health Expert Consensus Meeting - Part 1. Auton Neurosci 235:102828

69. Centers for Disease Control and Prevetion (2021) Symptoms and Diagnosis of ME/CFS. CDC

70. Bellach B-M, Ellert U, Radoschewski M (2000) Der SF-36 im Bundes-Gesundheitssurvey Erste Ergebnisse und neue Fragen: Erste Ergebnisse und neue Fragen. Bundesgesundheitsblatt-Gesundheitsforschung-Gesundheitsschutz 43:210–216

71. Jason L, Brown M, Evans M, Anderson V, Lerch A, Brown A, Hunnell J, Porter N (2011) Measuring substantial reductions in functioning in patients with chronic fatigue syndrome. Disabil Rehabil 33:589–598

72. Kristjánsdóttir J, Olsson GI, Sundelin C, Naessen T (2011) Could SF-36 be used as a screening instrument for depression in a Swedish youth population? Scandinavian journal of caring sciences 25:262–268

73. Office for National Statistics (2023) Prevalence of ongoing symptoms following coronavirus (COVID-19) infection in the UK: 30 March 2023.

74. Ludvigsson JF (2021) Case report and systematic review suggest that children may experience similar long-term effects to adults after clinical COVID-19. Acta Paediatr 110:914–921

75. Katz BZ, Shiraishi Y, Mears CJ, Binns HJ, Taylor R (2009) Chronic fatigue syndrome after infectious mononucleosis in adolescents. Pediatrics 124:189–193

76. Jason LA, Cotler J, Islam MF, Sunnquist M, Katz BZ (2021) Risks for Developing Myalgic Encephalomyelitis/Chronic Fatigue Syndrome in College Students Following Infectious Mononucleosis: A Prospective Cohort Study. Clin Infect Dis 73:e3740–e3746

77. Pricoco R, Meidel P, Hofberger T, Zietemann H, Mueller Y, Wiehler K, Michel K, Paulick J, Leone A, Haegele M, Mayer-Huber S, Gerrer K, Mittelstrass K, Scheibenbogen C, Renz-Polster H, Mihatsch L, Behrends U (2023) One-Year Follow-up of Young People with ME/CFS Following Infectious Mononucleosis by Epstein-Barr Virus. medRxiv:2023.2007.2024.23293082

78. Borch L, Holm M, Knudsen M, Ellermann-Eriksen S, Hagstroem S (2022) Long COVID symptoms and duration in SARS-CoV-2 positive children - a nationwide cohort study. Eur J Pediatr 181:1597–1607

79. VanElzakker MB, Brumfield SA, Lara Mejia PS (2018) Neuroinflammation and Cytokines in Myalgic Encephalomyelitis/Chronic Fatigue Syndrome (ME/CFS): A Critical Review of Research Methods. Front Neurol 9:1033

80. Conroy KE, Islam MF, Jason LA (2022) Evaluating case diagnostic criteria for myalgic encephalomyelitis/chronic fatigue syndrome (ME/CFS): toward an empirical case definition. Disability and Rehabilitation:1–8

81. Legler AF, Meyer-Arndt L, Mödl L, Kedor C, Freitag H, Stein E, Hoppmann U, Rust R, Konietschke F, Thiel A, Paul F, Scheibenbogen C, Bellmann-Strobl J (2023) Symptom persistence and biomarkers in post-COVID-19/chronic fatigue syndrome – results from a prospective observational cohort. medRxiv:2023.2004.2015.23288582

82. Sotzny F, Filgueiras IS, Kedor C, Freitag H, Wittke K, Bauer S, Sepulveda N, et al. (2022) Dysregulated autoantibodies targeting vaso- and immunoregulatory receptors in Post COVID Syndrome correlate with symptom severity. Front Immunol 13:981532

83. Haffke M, Freitag H, Rudolf G, Seifert M, Doehner W, Scherbakov N, Hanitsch L, Wittke K, Bauer S, Konietschke F, Paul F, Bellmann-Strobl J, Kedor C, Scheibenbogen C, Sotzny F (2022) Endothelial dysfunction and altered endothelial biomarkers in patients with post-COVID-19 syndrome and chronic fatigue syndrome (ME/CFS). J Transl Med 20:138

84. Flaskamp L, Roubal C, Uddin S, Sotzny F, Kedor C, Bauer S, Scheibenbogen C, Seifert M (2022) Serum of Post-COVID-19 Syndrome Patients with or without ME/CFS Differentially Affects Endothelial Cell Function In Vitro. Cells 11

85. Wong TL, Weitzer DJ (2021) Long COVID and Myalgic Encephalomyelitis/Chronic Fatigue Syndrome (ME/CFS)-A Systemic Review and Comparison of Clinical Presentation and Symptomatology. Medicina (Kaunas) 57

