## Supplementary material for "Pediatric and Adult Patients with ME/CFS following COVID-19: A Structured Approach to Diagnosis Using the Munich Berlin Symptom Questionnaire (MBSQ)": MBSQs and Supplementary Scoring Sheets

Klinikum rechts der Isar Technical University of Munich

CHARITÉ UNIVERSITÄTSMEDIZIN BERLIN

### Munich Berlin Symptom Questionnaire (MBSQ) – Questionnaire for Adults

| | |
|---|---|
| Surname: | Name (physician): |
| Name: | Date (physician): |
| Date of birth: | Institution: |
| Today's date: Completion time: min | Date of disease onset: |

*This questionnaire has to be used in **medical interview.** Open **questions** and comprehensive **problems** should be clarified in a **medical visit.** The medical **evaluation** of this questionnaire has to be based on the **supplementary scoring sheet for adults.** ME/CFS is a **clinical diagnosis.** The diagnosis cannot be established without appropriate **differential diagnostics.***

| | | During the last 6 months | | | |
|---|---|---|---|---|---|
| | | **Is not present** | **Frequency**<br>**1** = sometimes<br>**2** = about ½ of the time<br>**3** = most of the time<br>**4** = always | **Severity**<br>**1** = mild<br>**2** = moderate<br>**3** = severe<br>**4** = very severe | **Physician's notes** |
| **I** | **Fatigue/ Daily Function** | | | | |
| **1** | Fatigue (exhaustion, tiredness) | 0 | 1 2 3 4 | 1 2 3 4 | |
| **2** | Limitations in daily life | 0 | 1 2 3 4 | 1 2 3 4 | |

| | | |
|---|---|---|
| **3** | If fatigue is present, did it start new or at a definable time? | Yes No |
| **4** | If fatigue is present, is it the result of ongoing, excessive exertion? | Yes No |
| **5** | If fatigue is present, is it alleviated by rest? | Yes No |

| | | | | |
|---|---|---|---|---|
| **II** | **Post-Exertional Symptoms** | | | |
| **6** | Loss of physical or mental stamina | 0 | 1 2 3 4 | 1 2 3 4 |
| **7** | Increased symptoms after mild daily activities | 0 | 1 2 3 4 | 1 2 3 4 |

| | | |
|---|---|---|
| **8** | If post-exertional symptoms are present, how long does it take after mild daily activities until the increased symptoms have subsided? | O ≤ 1 h O 2-3 h O 4-10 h<br>O 11-13 h O 14-23 h O ≥ 24 h |
| **9** | If post-exertional symptoms are present, which three mild daily activities lead to a worsening of the symptoms? | 1.......................................<br>2.......................................<br>3....................................... |
| **10** | If post-exertional symptoms are present, which three symptoms worsen after mild daily activities? | 1.......................................<br>2.......................................<br>3....................................... |

| | | During the last 6 months | | | |
|---|---|---|---|---|---|
| | | **Not present** | **Frequency**<br>**1** = sometimes<br>**2** = about ½ of the time<br>**3** = most of the time<br>**4** = always | **Severity**<br>**1** = mild<br>**2** = moderate<br>**3** = severe<br>**4** = very severe | **Physician's notes** |
| **III** | **Sleep** | | | | |
| **11** | Unrefreshing sleep | 0 | 1 2 3 4 | 1 2 3 4 | |
| **12** | Unusually much sleep during the day | 0 | 1 2 3 4 | 1 2 3 4 | |
| **13** | Problems falling asleep | 0 | 1 2 3 4 | 1 2 3 4 | |
| **14** | Problems sleeping through | 0 | 1 2 3 4 | 1 2 3 4 | |
| **15** | Day / night reversal | 0 | 1 2 3 4 | 1 2 3 4 | |

| | | | | |
|---|---|---|---|---|
| **IV** | **Pain** | | | |
| **16** | Myofascial pain (can include achy and sore muscles) | 0 | 1 2 3 4 | 1 2 3 4 |
| **17** | Joint pain without swelling or redness | 0 | 1 2 3 4 | 1 2 3 4 |
| **18** | Headaches | 0 | 1 2 3 4 | 1 2 3 4 |

| | | | | |
|---|---|---|---|---|
| **V** | **Neurocognitive Manifestations** | | | |
| **19** | Confusion | 0 | 1 2 3 4 | 1 2 3 4 |
| **20** | Slowness of thought | 0 | 1 2 3 4 | 1 2 3 4 |
| **21** | Concentration problems | 0 | 1 2 3 4 | 1 2 3 4 |
| **22** | Memory problems | 0 | 1 2 3 4 | 1 2 3 4 |

| | | During the last 6 months | | | |
|---|---|---|---|---|---|
| | | Not present | **Frequency**<br>**1** = sometimes<br>**2** = about ½ of the time<br>**3** = most of the time<br>**4** = always | **Severity**<br>**1** = mild<br>**2** = moderate<br>**3** = severe<br>**4** = very severe | **Physician's notes** |
| **23** | Orientation problems | 0 | 1 2 3 4 | 1 2 3 4 | |
| **24** | Comprehension problems | 0 | 1 2 3 4 | 1 2 3 4 | |
| **25** | Word-finding problems | 0 | 1 2 3 4 | 1 2 3 4 | |
| **26** | If cognitive symptoms (**V.20-25**) are present, do they worsen due to effort / stress or time pressure? | | | Yes No | |
| **27** | Perceptual and sensory disturbances, e.g., blurred vision | 0 | 1 2 3 4 | 1 2 3 4 | |
| **28** | Coordination problems | 0 | 1 2 3 4 | 1 2 3 4 | |
| **29** | Muscle twitches | 0 | 1 2 3 4 | 1 2 3 4 | |
| **30** | Muscle weakness | 0 | 1 2 3 4 | 1 2 3 4 | |
| **31** | Overload phenomena - hypersensitivity to (bright) light | 0 | 1 2 3 4 | 1 2 3 4 | |
| **32** | Overload phenomena - hypersensitivity to noise | 0 | 1 2 3 4 | 1 2 3 4 | |
| **33** | Overload phenomena - hypersensitivity to touch | 0 | 1 2 3 4 | 1 2 3 4 | |
| **34** | Overload phenomena – emotional overload | 0 | 1 2 3 4 | 1 2 3 4 | |

| | | | | |
|---|---|---|---|---|
| **VI** | **Autonomic Manifestations** | | | |
| **35** | Dizziness | 0 | 1 2 3 4 | 1 2 3 4 |
| **36** | Palpitations with or without cardiac arrhythmias | 0 | 1 2 3 4 | 1 2 3 4 |
| **37** | Circulatory problems when getting up | 0 | 1 2 3 4 | 1 2 3 4 |
| **38** | Circulatory problems in an upright posture | 0 | 1 2 3 4 | 1 2 3 4 |
| **39** | Unusual paleness | 0 | 1 2 3 4 | 1 2 3 4 |
| **40** | Bladder problems | 0 | 1 2 3 4 | 1 2 3 4 |
| **41** | Gastrointestinal problems | 0 | 1 2 3 4 | 1 2 3 4 |
| **42** | Exertional shortness of breath | 0 | 1 2 3 4 | 1 2 3 4 |

| | | | | |
|---|---|---|---|---|
| **VII** | **Neuroendocrine Manifestations** | | | |
| **43** | Loss of thermostatic stability (e.g., subnormal body temperature, and marked diurnal fluctuation, sweating episodes, feelings of feverishness, cold limbs) | 0 | 1 2 3 4 | 1 2 3 4 |
| **44** | Intolerance of extremes hot and cold | 0 | 1 2 3 4 | 1 2 3 4 |
| **45** | Marked weight change – loss of appetite or abnormal appetite | 0 | 1 2 3 4 | 1 2 3 4 |
| **46** | Worsening of symptoms with stress | 0 | 1 2 3 4 | 1 2 3 4 |

| | | | | |
|---|---|---|---|---|
| **VIII** | **Immunologic Manifestations** | | | |
| **47** | Flu like symptoms / general malaise | 0 | 1 2 3 4 | 1 2 3 4 |
| **48** | Painful lymph nodes | 0 | 1 2 3 4 | 1 2 3 4 |
| **49** | Recurrent sore throat | 0 | 1 2 3 4 | 1 2 3 4 |
| **50** | New sensitivities to food, medication or chemicals | 0 | 1 2 3 4 | 1 2 3 4 |

| **Further Questions About the Symptom Course** | |
|---|---|
| If there are any symptoms,<br>what are the three main symptoms of this questionnaire? | 1……………………………………………………………………………<br>2……………………………………………………………………………<br>3…………………………………………………………………………… |

***For evaluation of this questionnaire please use the supplementary scoring sheet for adults.***

Klinikum rechts der Isar | Technical University of Munich | CHARITÉ UNIVERSITÄTSMEDIZIN BERLIN

### Munich Berlin Symptom Questionnaire (MBSQ) – Questionnaire for Pediatric and Adolescent Patients

| | |
|---|---|
| Surname: | Name (physician): |
| Name: | Date (physician): |
| Date of birth: | Institution: |
| Today's date: Completion time: min | Date of disease onset: |

*For patients: Please complete this questionnaire on your own as far as possible. If necessary, ask your parents to help you.*
*For doctors: This questionnaire has to be used in **medical interview.** Open **questions** and comprehensive **problems** should be clarified in a **medical visit.** The medical **evaluation** of this questionnaire has to be based on the **supplementary scoring sheet for adults.** ME/CFS is a **clinical diagnosis.** The diagnosis cannot be established without appropriate **differential diagnostics.***

| | | During the last 3 months | | | |
|---|---|---|---|---|---|
| | | **Is not present** | **Frequency**<br>**1** = sometimes<br>**2** = about ½ of the time<br>**3** = most of the time<br>**4** = always | **Severity**<br>**1** = mild<br>**2** = moderate<br>**3** = severe<br>**4** = very severe | **Physician's notes** |
| **I** | **Fatigue / Daily Function** | | | | |
| **1** | Fatigue (exhaustion, tiredness) | 0 | 1 2 3 4 | 1 2 3 4 | |
| **2** | Limitations in daily life - school/ education | 0 | 1 2 3 4 | 1 2 3 4 | |
| **3** | Limitations in daily life - social | 0 | 1 2 3 4 | 1 2 3 4 | |
| **4** | Limitations in daily life - personal | 0 | 1 2 3 4 | 1 2 3 4 | |

| | | |
|---|---|---|
| **5** | If fatigue is present, did it start new or at a definable time? | Yes No |
| **6** | If fatigue is present, is it the result of ongoing, excessive exertion? | Yes No |
| **7** | If fatigue is present, is it alleviated by rest? | Yes No |

| | | | | |
|---|---|---|---|---|
| **II** | **Post-Exertional Symptoms** | | | |
| **8** | Loss of physical or mental stamina | 0 | 1 2 3 4 | 1 2 3 4 |
| **9** | Increased symptoms after light everyday activities | 0 | 1 2 3 4 | 1 2 3 4 |

| | | |
|---|---|---|
| **10** | If post-exertional symptoms are present, how long does it take after mild daily activities until the increased symptoms have subsided? | O ≤ 1 h O 2-3 h O 4-10 h<br>O 11-13 h O 14-23 h O ≥ 24 h |
| **11** | If post-exertional symptoms are present, which three mild daily activities lead to a worsening of the symptoms? | 1.......................................<br>2.......................................<br>3....................................... |
| **12** | If post-exertional symptoms are present, which three symptoms worsen after mild daily activities? | 1.......................................<br>2.......................................<br>3....................................... |

| | | During the last 3 months | | | |
|---|---|---|---|---|---|
| | | **Not present** | **Frequency**<br>**1** = sometimes<br>**2** = about ½ of the time<br>**3** = most of the time<br>**4** = always | **Severity**<br>**1** = mild<br>**2** = moderate<br>**3** = severe<br>**4** = very severe | **Physician's notes** |
| **III** | **Sleep** | | | | |
| **13** | Unrefreshing sleep | 0 | 1 2 3 4 | 1 2 3 4 | |
| **14** | Unusually much sleep during the day | 0 | 1 2 3 4 | 1 2 3 4 | |
| **15** | Problems falling asleep | 0 | 1 2 3 4 | 1 2 3 4 | |
| **16** | Problems sleeping through | 0 | 1 2 3 4 | 1 2 3 4 | |
| **17** | Day/ night reversal | 0 | 1 2 3 4 | 1 2 3 4 | |

| | | | | |
|---|---|---|---|---|
| **IV** | **Pain** | | | |
| **18** | Myofascial pain (can include achy and sore muscles) | 0 | 1 2 3 4 | 1 2 3 4 |
| **19** | Joint pain without swelling or redness | 0 | 1 2 3 4 | 1 2 3 4 |
| **20** | Headaches | 0 | 1 2 3 4 | 1 2 3 4 |
| **21** | Abdominal pain | 0 | 1 2 3 4 | 1 2 3 4 |

| | During the last 3 months | | | |
|---|---|---|---|---|
| | Not present | **Frequency**<br>**1** = sometimes<br>**2** = about ½ of the time<br>**3** = most of the time<br>**4** = always | **Severity**<br>**1** = mild<br>**2** = moderate<br>**3** = severe<br>**4** = very severe | **Physician's notes** |
| **V Neurocognitive Manifestations** | | | | |
| **22** Confusion | 0 | 1 2 3 4 | 1 2 3 4 | |
| **23** Slowness of thought | 0 | 1 2 3 4 | 1 2 3 4 | |
| **24** Concentration problems | 0 | 1 2 3 4 | 1 2 3 4 | |
| **25** Memory problems | 0 | 1 2 3 4 | 1 2 3 4 | |
| **26** Orientation problems | 0 | 1 2 3 4 | 1 2 3 4 | |
| **27** Comprehension problems | 0 | 1 2 3 4 | 1 2 3 4 | |
| **28** Word-finding problems | 0 | 1 2 3 4 | 1 2 3 4 | |

| | |
|---|---|
| **29** If cognitive symptoms (**V.23-28**) are present, do they worsen due to Effort / stress or time pressure? | Yes No |

| | | | |
|---|---|---|---|
| **30** Absent mindedness | 0 | 1 2 3 4 | 1 2 3 4 |
| **31** Difficulty recalling information | 0 | 1 2 3 4 | 1 2 3 4 |
| **32** Need to focus on one thing at a time | 0 | 1 2 3 4 | 1 2 3 4 |
| **33** Trouble expressing thought | 0 | 1 2 3 4 | 1 2 3 4 |
| **34** Lose train of thought | 0 | 1 2 3 4 | 1 2 3 4 |
| **35** New troubles with math or other educational subjects | 0 | 1 2 3 4 | 1 2 3 4 |
| **36** Perceptual and sensory disturbances, e.g., blurred vision | 0 | 1 2 3 4 | 1 2 3 4 |
| **37** Coordination problems | 0 | 1 2 3 4 | 1 2 3 4 |
| **38** Muscle twitches | 0 | 1 2 3 4 | 1 2 3 4 |
| **39** Muscle weakness | 0 | 1 2 3 4 | 1 2 3 4 |
| **40** Overload phenomena - hypersensitivity to (bright) light | 0 | 1 2 3 4 | 1 2 3 4 |
| **41** Overload phenomena - hypersensitivity to noise | 0 | 1 2 3 4 | 1 2 3 4 |
| **42** Overload phenomena - hypersensitivity to touch | 0 | 1 2 3 4 | 1 2 3 4 |
| **43** Overload phenomena – emotional overload | 0 | 1 2 3 4 | 1 2 3 4 |

| | | | |
|---|---|---|---|
| **VI Autonomic Manifestations** | | | |
| **44** Dizziness | 0 | 1 2 3 4 | 1 2 3 4 |
| **45** Palpitations with or without cardiac arrhythmias | 0 | 1 2 3 4 | 1 2 3 4 |
| **46** Circulatory problems when getting up | 0 | 1 2 3 4 | 1 2 3 4 |
| **47** Circulatory problems in upright posture | 0 | 1 2 3 4 | 1 2 3 4 |
| **48** Unusual paleness | 0 | 1 2 3 4 | 1 2 3 4 |
| **49** Bladder problems | 0 | 1 2 3 4 | 1 2 3 4 |
| **50** Gastrointestinal problems | 0 | 1 2 3 4 | 1 2 3 4 |
| **51** Exertional shortness of breath | 0 | 1 2 3 4 | 1 2 3 4 |

| | | | |
|---|---|---|---|
| **VII Neuroendocrine Manifestations** | | | |
| **52** Loss of thermostatic stability (e.g., subnormal body temperature, and marked diurnal fluctuation, sweating episodes, feelings of feverishness, cold limbs) | 0 | 1 2 3 4 | 1 2 3 4 |
| **53** Intolerance of extremes hot and cold | 0 | 1 2 3 4 | 1 2 3 4 |
| **54** Marked weight change – loss of appetite or abnormal appetite | 0 | 1 2 3 4 | 1 2 3 4 |
| **55** Worsening of symptoms with stress | 0 | 1 2 3 4 | 1 2 3 4 |

| | | | |
|---|---|---|---|
| **VIII Immunologic Manifestations** | | | |
| **56** Flu like symptoms / general malaise | 0 | 1 2 3 4 | 1 2 3 4 |
| **57** Painful lymph nodes | 0 | 1 2 3 4 | 1 2 3 4 |
| **58** Fever | 0 | 1 2 3 4 | 1 2 3 4 |
| **59** Recurrent sore throat | 0 | 1 2 3 4 | 1 2 3 4 |
| **60** New sensitivities to food, medication or chemicals | 0 | 1 2 3 4 | 1 2 3 4 |

| | |
|---|---|
| **Further Questions About the Symptom Course** | |
| If there are any symptoms,<br>what are the three main symptoms of this questionnaire? | 1……………………………………………………………………………<br>2……………………………………………………………………………<br>3…………………………………………………………………………… |

Klinikum rechts der Isar Technical University of Munich

CHARITÉ UNIVERSITÄTSMEDIZIN BERLIN

### Munich Berlin Symptom Questionnaire – Scoring Sheet for Adults (≥ 18 Years of Age)

| | |
|---|---|
| Surname: | Surname (physician): |
| Name: | Name (physician): |
| Date of birth: | Date (physician): |
| Today's date: | Institution: |

***Only if frequency and severity are reported with ≥2, the respective item counts positively for evaluation.***

| | Canadian Consensus Criteria[1] | | IOM Criteria[2] |
|---|---|---|---|
| O | **Duration of Illness (Onset of the Symptoms: ____.____.________)**<br>The symptomatology is present for at least **6 months**. | O | **Duration of Illness (Onset of the Symptoms: ____.____.________)**<br>The symptomatology is present for at least **6 months.** |
| O | **Medical History, Physical Examination, and Differential Diagnostics**<br>☐ did not indicate any other cause for the symptoms<br>*(in particular, no indications for Addison's disease, Cushing's disease, hypo-/hyperthyroidism, anemia, hemochromatosis, diabetes mellitus, haemato-oncological, rheumatological and treatable sleep disorders)*<br>☐ The symptoms must have begun or have been significantly altered after the onset of the illness. | O | **Medical History, Physical Examination, and Differential Diagnostics**<br>☐ did not indicate any other cause for the symptoms<br>☐ The symptoms must have begun or have been significantly altered after the onset of the illness. |
| O | **Fatigue / Daily Function**<br>All of the following points must apply:<br>☐ Fatigue: **I.1** ≥2<br>☐ Limitations: at least 1 of the 3 points **I.2** ≥2<br>☐ Fatigue new start: **I.3** Yes | O | **Fatigue / Daily Function**<br>All of the following points must apply:<br>☐ Fatigue: **I.1** ≥2<br>☐ Limitations: at least 1 of the 3 points **I.2** ≥2<br>☐ Fatigue new start: **I.3** Yes<br>☐ Fatigue due to exertion: **I.4** No<br>☐ Fatigue alleviated by rest: **I.5** No |
| O | **Post-Exertional Symptoms**<br>All of the following points must apply:<br>☐ loss of stamina; increased symptoms after everyday activities: all 2 points **II.6-7** ≥2<br>☐ duration of worsening of symptoms: **II.8** ≥14 hours* | O | **Post-Exertional Symptoms**<br>All of the following points must apply: all 2 points **II.6-7** ≥2 |
| O | **Sleep**<br>At least 1 of the following 5 points must apply: **III.11-15** ≥2 | O | **Sleep**<br>The following point must apply: **III.11** ≥2 |

| Canadian Consensus Criteria[1] | | IOM Criteria[2] | |
|---|---|---|---|
| O | **Pain**<br>At least 1 of the following 5 points must apply: **VI.16-18** ≥2 | | **Pain**<br>*Not included in IOM criteria* |
| O | **Neurocognitive Manifestations**<br>At least 2 of the following symptom groups must apply:<br>□ Confusion: **V.19** ≥2<br>□ Concentration and/or Memory: 1 of the following 2 points: **V.21-22** ≥2<br>□ Orientation: **V.23** ≥2<br>□ Comprehension and/or word-finding: 1 of the following 2 points: **V.24-25** ≥2<br>□ Perception and overload phenomena: 1 of the following 5 points: **V.27, V.31-34** ≥2<br>□ Coordination and muscular system: 1 of the following 3 points: **V.28-30** ≥2 | O | **Neurocognitive and Autonomous Manifestations**<br>At least 1 of the following 2 categories must apply:<br>□ The 2 following symptom groups must apply:<br>□ At least 1 of the following 6 points must apply: **V.20-22, V.24-25, V.28** ≥2<br>□ The following point must apply: **V.26** Yes<br>□ At least 1 of the following 2 points must apply: **VI.37-38** ≥2 |
| O | **Autonomic, Neuroendocrine, Immunologic Manifestations**<br>At least 2 of the following 3 categories must apply:<br>□ At least 1 of the following 8 points must apply: **VI.35-42** ≥2<br>□ At least 1 of the following 4 points must apply: **VII.43-46** ≥2<br>□ At least 1 of the following 4 points must apply: **VIII.47-50**, ≥2 | | **Neuroendocrine, Immunologic Manifestations**<br>*Not included in IOM criteria* |

| | |
|---|---|
| O **Patient meets Canadian Consensus Criteria for ME/CFS**<br>O **Patient does not meet Canadian Consensus Criteria for ME/CFS** | O **Patient meets IOM criteria for ME/CFS**<br>O **Patient does not meet IOM criteria for ME/CFS** |

***In discussion with the patient & in the physician's letter, it must be pointed out that ME/CFS is a diagnosis of exclusion, which must be re-evaluated in case of new clinical aspects.***

[1] Carruthers, Bruce & Jain, Anil & Meirleir, Kenny & Peterson, Daniel & Klimas, Nancy & Lerner, A. & Bested, Alison & Flor-Henry, Pierre & Joshi, Pradip & Powles, Peter & Sherkey, Jeffrey & Sande, Marjorie. (2003). Myalgic Encephalomyelitis/Chronic Fatigue Syndrome. Journal Of Chronic Fatigue Syndrome. 11. 7-115. 10.1300/J092v11n01_02.

[2] Clayton, Ellen. (2015). Beyond Myalgic Encephalomyelitis/Chronic Fatigue Syndrome An IOM Report on Redefining an Illness. JAMA. 313. 10.1001/jama.2015.1346.

* In the original publication of Carruthers et al.[1] PEM requires a duration of >24 hours. However, according to the current state of international research, we recommend diagnosing PEM with a duration of ≥14 hours (see Cotler J et al. (2018) A Brief Questionnaire to Assess Post-Exertional Malaise. Diagnostics (Basel). 2018;8(3):66. Doi: 10.3390/diagnostics8030066 und Kedor, C., Freitag, H., Meyer-Arndt, L. et al. A prospective observational study of post-COVID-19 chronic fatigue syndrome following the first pandemic wave in Germany and biomarkers associated with symptom severity. Nat Commun 13, 5104 (2022). https://doi.org/10.1038/s41467-022-32507-6).

Klinikum rechts der Isar    Technical University of Munich

CHARITÉ UNIVERSITÄTSMEDIZIN BERLIN

### Munich Berlin Symptom Questionnaire – Scoring Sheet for Children and Adolescents (0-17 Years of Age)

| | |
|---|---|
| Surname: | Surname (physician): |
| Name: | Name (physician): |
| Date of birth: | Date (physician): |
| Today's date: | Institution: |

***Only if frequency and severity are reported with ≥2, the respective item counts positively for evaluation.***

| | Pediatric Case Definition for ME/CFS by Jason et al.[1] | | Clinical Diagnostic Worksheet by Rowe et al.[2] |
|---|---|---|---|
| O | **Duration of Illness (Onset of the Symptoms: ____.____.________)**<br>The symptomatology is present for at least **3 months**. | O | **Duration of Illness (Onset of the Symptoms: ____.____.________)**<br>The symptomatology is present for at least **3 months**. |
| O | **Medical History, Physical Examination and Differential Diagnostics**<br>did not indicate any other cause for the symptoms<br>*(in particular, no indications for untreated hypothyreoidism, sleep apnea, narcolepsy, malignancies, leukemia, unresolved hepatitis, multiple sclerosis, juvenile rheumatoid arthritis, lupus erythematosus, HIV/AIDS, severe obesity (BMI greater than 40), celiac disease, lyme disease, childhood schizophrenia or psychiatric disorders, bipolar disorder, active alcohol or substance abuse, active anorexia nervosa or bulimia nervosa, depressive disorders)* | O | **Medical History, Physical Examination and Differential Diagnostics**<br>did not indicate any other cause for the symptoms<br>*(in particular, no evidence of adrenocortical insufficiency, overtraining syndrome, GI disorders: Celiac disease, irritable bowel syndrome, eosinophilic gastroenteritis; Chiari malformation, cervical spinal stenosis, neuroborreliosis or other tick-borne disease, major depression, narcolepsy, obstructive or central sleep apnea, postcommotion syndrome, severe anemia, systemic lupus erythematosus and similar autoimmune disease, untreated hypo-/hyperthyroidism).* |
| O | **Fatigue / Daily Function**<br>All of the following points must apply:<br>☐ Fatigue: **I.1** ≥2<br>☐ Limitations: all 3 points: **I.2-4** ≥2<br>☐ Fatigue due to exertion: **I.6** No<br>☐ Fatigue alleviated by rest: **I.7** No | O | **Fatigue / Daily Function**<br>All of the following points must apply:<br>☐ Fatigue: **I.1** ≥2<br>☐ Limitations: at least 1 of the 3 points: **I.2-4** ≥2<br>☐ Fatigue due to exertion: **I.6** No<br>☐ Fatigue alleviated by rest: **I.7** No |
| O | **Post-Exertional Symptoms**<br>All of the following points must apply:<br>☐ loss of stamina; increased symptoms after everyday activities: all 2 points: **II.8-9** ≥2<br>☐ duration of worsening of symptoms: **II.10** ≥14 hours | O | **Post-Exertional Symptoms**<br>All of the following points must apply:<br>☐ loss of stamina; increased symptoms after everyday activities: all 2 points: **II.8-9** ≥2<br>☐ duration of worsening of symptoms: **II.10** >24 hours |

Klinikum rechts der Isar   Technical University of Munich

CHARITÉ UNIVERSITÄTSMEDIZIN BERLIN

| Pediatric Case Definition for ME/CFS by Jason et al.[1] | | Clinical Diagnostic Worksheet by Rowe et al.[2] | |
|---|---|---|---|
| O | **Sleep**<br>At least 1 of the following 5 points must apply: **III.13-17** ≥2 | O | **Sleep, Pain, and Neurocognitive Symptoms**<br>At least 2 of the following 3 categories must apply:<br>☐ At least 1 of the following 5 points must apply: **III.13-17** ≥2<br>☐ At least 1 of the following 6 points must apply: **IV.18-21, IV.57, IV.59** ≥2<br>☐ At least 1 of the following 8 points must apply: **V.23-25, V.27-28, V.30, V.33** ≥2; **V.29**: Yes |
| O | **Pain**<br>At least 1 of the following 4 points must apply: **IV.18-21** ≥2 | | |
| O | **Neurocognitive Manifestations**<br>At least 2 of the following 11 points must apply: **V.23-25; V.27-28; V.30-35** ≥2 | | |
| O | **Autonomic, Neuroendocrine and Immunologic Manifestations**<br>At least 2 of the following 3 categories must apply:<br>☐ At least 1 of the following 5 points must apply: **VI.44-47; VI.51** ≥2<br>☐ At least 1 of the following 4 points must apply: **VII.52-55** ≥2<br>☐ At least 1 of the following 4 points must apply: **VIII.56-59** ≥2 | O | **Autonomic, Neuroendocrine, and Immunologic Manifestations**<br>*Not included in the Clinical Diagnostic Worksheet by Rowe* |

| | |
|---|---|
| O **Patient meets the Pediatric Case Definition for ME/CFS by Jason** | O **Patient meets the Clinical Diagnostic Worksheet criteria for ME/CFS by Rowe** |
| O **Patient does not meet the Pediatric Case Definition for ME/CFS by Jason** | O **Patient does not meet the Clinical Diagnostic Worksheet criteria for ME/CFS by Rowe** |

***In the discussion with patients and parents as well as in the physician's letter it must be pointed out that ME/CFS is clinical diagnosis which must be re-evaluated in the case of new clinical aspects.***

[1]Jason, Leonard & Jordan, Karen & Miike, Teruhisa & Bell, David & Lapp, Charles & Torres-Harding, Susan & Rowe, Kathy & Gurwitt, Alan & Meirleir, Kenny & Hoof, Elke & Psych, Clin & Chairperson,. (2006). A Pediatric Case Definition for Myalgic Encephalomyelitis and Chronic Fatigue Syndrome. Journal Of Chronic Fatigue Syndrome. 13. 1-44. 10.1300/J092v13n02_01.

[2] Rowe, Peter & Underhill, Rosemary & Friedman, Kenneth & Gurwitt, Alan & Medow, Marvin & Schwartz, Malcolm & Speight, Nigel & Stewart, Julian & Vallings, Rosamund & Rowe, Katherine. (2017). Myalgic Encephalomyelitis/Chronic Fatigue Syndrome Diagnosis and Management in Young People: A Primer. Frontiers in Pediatrics. 5. 10.3389/fped.2017.00121.

* In the original publication by Rowe et al.[2] a duration of at least 6 months is required for the diagnosis. If the duration of the disease is <6 months, it is only recommended to express an urgent suspicion. We recommend a diagnosis in children and adolescents after a duration of 3 months in order to provide symptom-oriented care at an early stage.

Klinikum rechts der Isar Technical University of Munich

CHARITÉ UNIVERSITÄTSMEDIZIN BERLIN

### Munich Berlin Symptom Questionnaire – Additional Scoring Sheet for Adolescents (12-17 Years)

***Only if frequency and severity are reported with ≥2, the respective item enters positively into the evaluation.***

| | |
|---|---|
| Surname: | Surname (physician): |
| Name: | Name (physician): |
| Date of birth: | Date (physician): |
| Today's date: | Institution: |

| | Canadian Consensus Criteria[3] | | IOM Criteria[4] |
|---|---|---|---|
| O | **Duration of Illness (Onset of the Symptoms: ____.____.________)**<br>The symptomatology has been present for at least **3 months**. | O | **Duration of Illness (Onset of the Symptoms: ____.____.________)**<br>The symptomatology has been present for at least **3 months**.* |
| O | **Medical History, Physical Examination and Differential Diagnostics**<br>☐ did not indicate any other cause for the symptoms *(in particular no indications for Addison's disease, Cushing's disease, hypo-/hyperthyroidism, anemia, hemochromatosis, diabetes mellitus, haemato-oncological, rheumatological and treatable sleep disorders)*<br>☐ The symptoms must have begun or have been significantly altered after the onset of the illness. | O | **Medical History, Physical Examination and Differential Diagnostics**<br>☐ did not indicate any other cause for the symptoms<br>☐ The symptoms must have begun or have been significantly altered after the onset of the illness. |
| O | **Fatigue / Daily Function**<br>All of the following points must apply:<br>☐ Fatigue: **I.1** ≥2<br>☐ Limitations: at least 1 of the 3 points: **I.2-4** ≥2<br>☐ Fatigue new start: **I.5** Yes | O | **Fatigue / Daily Function**<br>All of the following points must apply:<br>☐ Fatigue: **I.1** ≥2<br>☐ Limitations: at least 1 of the 3 points: **I.2-4** ≥2<br>☐ Fatigue new start: **I.5** Yes<br>☐ Fatigue due to exertion: **I.6** No<br>☐ Fatigue alleviated by rest: **I.7** No |
| O | **Post-Exertional Symptoms**<br>All of the following points must apply:<br>☐ loss of stamina; increased symptoms after everyday activities: all 2 points **II.8-9** ≥2<br>☐ duration of worsening of symptoms: **II.10** >24 hours | O | **Post-Exertional Symptoms**<br>All of the following points must apply: all 2 points: **II.8-9** ≥2 |

| Canadian Consensus Criteria[3] | IOM Criteria[4] |
|---|---|
| O **Sleep**<br>At least 1 of the following 5 points must apply: **III.13-17** ≥2 | O **Sleep**<br>The following point must apply: **III.13** ≥2 |
| O **Pain**<br>At least 1 of the following 3 points must apply: **IV.18-20** ≥2<br>*(except abdominal pain)* | O **Pain**<br>*Not included in IOM criteria* |
| O **Neurocognitive Manifestations**<br>At least 2 of the following symptom groups must apply:<br>□ Confusion: **V.22** ≥2<br>□ Concentration and/or Memory: 1 of the following 2 points: **V.24-25** ≥2<br>□ Orientation: **V.26** ≥2<br>□ Comprehension and/or word-finding: 1 of the following 2 points: **V.27-28** ≥2<br>□ Perception and overload phenomena: 1 of the following 5 points: **V.36**, **V.40-43** ≥2<br>□ Coordination und muscular system: 1 of the following 3 points: **V.37-39** ≥2 | O **Neurocognitive and Autonomic Manifestations**<br>At least 1 of the following 2 categories must apply:<br>□ The 2 following symptom groups must apply:<br>□ At least 1 of the following 6 points must apply: **V.23-25, V.27-28**, **V.37** ≥2<br>□ The following point must apply: **V.29**, Yes<br>□ At least 1 of the following 2 points must apply: **VI.46-47** ≥2 |
| O **Autonomic, Neuroendocrine, Immunologic Manifestations**<br>At least 2 of the following 3 categories must apply:<br>□ At least 1 of the following 8 points must apply: **VI.44-51** ≥2<br>□ At least 1 of the following 4 points must apply: **VII.52-55** ≥2<br>□ At least 1 of the following 4 points must apply: **VIII.56-57, VIII.59-60** ≥2 | O **Neuroendocrine, Immunologic Manifestations**<br>*Not included in IOM criteria* |

| | |
|---|---|
| O **Patient meets Canadian Consensus Criteria for ME/CFS** | O **Patient meets IOM criteria for ME/CFS** |
| O **Patient does not meet Canadian Consensus Criteria for ME/CFS** | O **Patient does not meet IOM criteria for ME/CFS** |

***In the discussion with patients and parents as well as in the physician's letter it must be pointed out that ME/CFS is clinical diagnosis which must be re-evaluated in the case of new clinical aspects.***

[3] Carruthers, Bruce & Jain, Anil & Meirleir, Kenny & Peterson, Daniel & Klimas, Nancy & Lerner, A. & Bested, Alison & Flor-Henry, Pierre & Joshi, Pradip & Powles, Peter & Sherkey, Jeffrey & Sande, Marjorie. (2003). Myalgic Encephalomyelitis/Chronic Fatigue Syndrome. Journal Of Chronic Fatigue Syndrome. 11. 7-115. 10.1300/J092v11n01_02.

[4] Clayton, Ellen. (2015). Beyond Myalgic Encephalomyelitis/Chronic Fatigue Syndrome An IOM Report on Redefining an Illness. JAMA. 313. 10.1001/jama.2015.1346.

* In the original publication by Clayton et al.[4] a duration of at least 6 months is required for the diagnosis. We recommend diagnosing children and adolescents after a disease duration of 3 months to provide them with symptom-oriented care at an early stage.
